# Evaluation of an in vivo biomarker of arteriolosclerosis (ARTS) and its associations with cognition and multimodal ATN(V) biomarkers in a cardiometabolic-risk enriched community cohort

**DOI:** 10.64898/2025.12.23.25342940

**Authors:** Marc D. Rudolph, Samuel N. Lockhart, Melissa R. Rundle, Richard A. Barcus, Kathryn A. Alphin, James R. Bateman, Kiran K. Solingapuram Sai, Michelle M. Mielke, Thomas C. Register, Suzanne Craft, Shannon L. Risacher, Timothy M. Hughes

**Affiliations:** Department of Internal Medicine, Section on Gerontology and Geriatric Medicine, Wake Forest University School of Medicine, Winston-Salem, NC, USA; Department of Neurology, Virginia Commonwealth University, Richmond, VA, USA; Department of Radiology, Wake Forest University School of Medicine, Winston-Salem, NC, USA; Department of Epidemiology and Prevention, Wake Forest University School of Medicine, Winston-Salem, NC, USA; Department of Pathology, Section on Comparative Medicine, Wake Forest University School of Medicine, Winston-Salem, North Carolina, USA

**Author notes:** **Corresponding authors:** Marc D. Rudolph, PhD and Timothy M. Hughes, PhD Wake Forest School of Medicine, Medical Center Blvd, Winston-Salem, NC 27157.

## Abstract

**Objective:** To evaluate associations between an *in vivo* (MRI) marker of arteriolosclerosis (ARTS) and multimodal neuroimaging and plasma ATN(V) biomarkers.

**Methods:** Among 238 participants with both amyloid and tau PET scans within one year of MRI, we examined multivariable adjusted models relating ARTS with structural MRI (cortical thickness/volume, white matter hyperintensities [WMH]), diffusion MRI (fractional anisotropy [FA], mean diffusivity [MD], NODDI free water [FW]), cerebral blood flow, plasma biomarkers (p-tau217, A*β*42/40, neurofilament light, glial fibrillary acidic protein [GFAP]), and PET imaging.

**Results:** As expected, ARTS was most strongly linked to age and greater WMH burden and diffusion-based indices of microstructural disruption (FA, MD, FW). ARTS was elevated in ATN biomarker-positive groups (highest in A+T+N+) and was associated with greater neurodegeneration and higher plasma biomarker levels, GFAP in particular.

**Conclusions:** ARTS relates to other markers of vascular brain injury, neurodegeneration, amyloid and tau pathology within the ATN(V) framework, and inflammation.

**Highlights:**

- ARTS scores are elevated in ATN-positive individuals, most prominently in A+T+N+
- ARTS may exert stage-specific effects on neurodegeneration rather than track A/T burden.
- Strong ARTS-GFAP association suggests role of astroglial activation in vascular-related neurodegeneration
- Findings underscore the importance of vascular contributions to AD/ADRD pathophysiology, especially in high-cardiometabolic-risk populations

## 1 Introduction

Alzheimer’s disease and related dementias (AD/ADRD) are increasingly recognized as multifactorial disorders in which vascular injury often plays a role alongside amyloid and tau pathology. Approximately 70–80% of dementia cases present with mixed etiology, the majority exhibiting concomitant vascular pathology that often emerges in midlife.^1,2^ The revised ATN(V) framework for AD, encompassing amyloid (A), tau (T), neurodegeneration (N), *and* vascular injury (V), provides a unifying structure for integrating these processes into a single biomarker model. Cerebral small-vessel disease (cSVD) frequently co-occurs with AD/ADRD pathology^3–5^ and is increasingly recognized as a contributor to AD/ADRD.^6,7^

cSVD represents various forms of vascular pathology visible on brain MRI^8^ as well as microstructural changes not visible on imaging; therefore, multiple (V) biomarkers likely need to be evaluated within the context of the revised ATN(V) framework. Arteriolosclerosis, hardening and loss of elasticity of arterioles or small arteries, is a hallmark of cSVD frequently associated with neurodegeneration, impaired cognitive functioning^9^ and cognitive decline,^10^ and increased odds of AD.^11^ Tau and amyloid pathology may potentiate the microinfarct burden that characterizes cSVD,^4^ highlighting a synergistic interaction between vascular injury and neurodegenerative processes. Despite growing post-mortem evidence suggesting associations of arteriolosclerosis with AD pathology, *in vivo* measurement remains challenging.^12^ Traditional MRI markers of cSVD are nonspecific, indirect proxies that lack specificity for underlying arteriolosclerosis pathology, particularly when considered in isolation.

To advance in vivo measurement of arteriolosclerosis and increase understanding of the vascular contributions to cognitive impairment and dementia (VCID), the MarkVCID consortium developed the ARTS classifier, a machine learning-based tool trained on ex vivo MRI and neuropathology data and validated across three independent cohorts.^12^ The ARTS toolkit developed by the MarkVCID consortium offers a semi-automated resource for indexing arteriolosclerosis *in vivo* across the research community. While ARTS represents a major methodological advancement, its relationship to AD/ADRD biomarkers, particularly within the ATN(V) framework, remains underexplored. Two prior studies reported that ARTS was associated with peripheral vascular risk factors, small-vessel injury on MRI, and cognitive outcomes.^13,14^ However, studies have not examined ARTS within the ATN(V) framework using amyloid/tau PET or plasma biomarkers such as phosphorylated tau 217 (p-tau217), neurofilament light chain (NfL), or glial fibrillary acidic protein (GFAP), limiting insight into the biological mechanisms underlying these associations.

To address this gap, we investigated ARTS in relation to multimodal imaging (primary) and plasma (secondary) biomarkers within the ATN(V) framework^15^ in a deeply phenotyped, community-based cohort of older adults enrolled in the Clinical Core of the Wake Forest Alzheimer’s Disease Research Center (WF ADRC). As described previously, the WF ADRC cohort is characterized by elevated rates of obesity, hypertension, and cardiovascular disease relative to national averages,^16^ making it well-suited for establishing relationships between ARTS and AD/ADRD pathology in the context of cardiometabolic disorders. All included participants had MRI, amyloid, *and* tau PET; a subset of these participants had plasma biomarkers. Tau PET imaging, in particular, a key component of the ATN framework,^15^ offers a unique opportunity to contextualize vascular contributions within the broader landscape of AD/ADRD biomarkers given its established role as a robust predictor of clinical progression and its ability to stratify individuals by risk.^17–19^ We hypothesized that higher ARTS scores would be associated with greater amyloid and tau burden and altered plasma biomarker profiles, reflecting the vascular contributions to AD/ADRD pathology within the ATN(V) framework. To our knowledge, this is the first study to integrate ARTS with amyloid/tau PET and plasma biomarkers within the ATN(V) framework in a high-cardiometabolic-risk, community-based cohort.

## 2 METHODS

### 2.1 Participants

Participants between the ages of 55 and 85 were recruited from the surrounding community into the WF ADRC via the WF ADRC Clinical Core supported by efforts of the Outreach, Engagement & Recruitment Core.^20,21^ No participants received or were recruited with the intent of receiving or being recommended for disease-modifying treatments or therapies, nor were participants referred to our study for treatment (previously published exclusionary criteria are provided in the supplement for reference). Participants underwent a standard evaluation, including the National Alzheimer’s Coordinating Center (NACC) protocol for clinical research data collection, clinical exams, neurocognitive testing, neuroimaging (MRI, amyloid PET, and tau PET), blood collection for plasma-based biomarkers, and genotyping for the apolipoprotein E (*APOE*) ε4 allele. The Wake Forest Institutional Review Board approved all activities as described; written informed consent was obtained for all participants and/or their legally authorized representatives.

### 2.2 Clinical and Cognitive evaluation

An expert panel of investigators including neuropsychologists, neurologists, and geriatricians provided adjudication of cognitive status in accordance with current National Institute of Aging-Alzheimer’s Association guidelines. Following determination of clinical cognitive diagnosis for cognitively unimpaired (CU), mild cognitive impairment (MCI),^22^ and dementia (DEM),^23^ adjudication of cause/type of cognitive impairment or dementia was assessed using neuroimaging and fluid biomarkers. Cognition was assessed using a modified Preclinical Alzheimer’s Cognitive Composite (mPACC5) comprised of scores obtained from in-person visits on the MMSE, FCSRT, Craft Story delayed verbatim recall, Digit Symbol Substitution Test, and category fluency assessments.^24,25^

### 2.3 Hypertension and Impaired Glucose Tolerance

Brachial blood pressure was measured in a seated position after a 5-minute rest in a quiet room as described previously,^20^ and was categorized according to 2017 ACC/AHA guidelines,^26^ with hypertension (HTN) status defined as systolic blood pressure (SBP) ≥ 140 mmHg and/or having a history HTN or current use of antihypertensive medications as indicated on UDSv3 form *d1*. Approximately 63% of the analytical sample reported current or recent use hypertensive medications (CU = 59% [*n* =78]; MCI = 70% [*n*=56]; DEM = 54% [*n* =14]; ∼84% of those classified as having hypertension (as described above). Irrespective of HTN status, 76% of those with high blood pressure, as defined here, reported hypertensive medication use (18% with uncontrolled blood pressure). Fasting blood glucose levels were measured via an oral glucose tolerance test (OGTT) from serial blood draws; impaired glucose tolerance (IGT) was defined as fasting glucose tolerance > 100 mg/dL or hemoglobin A1c > 5.6%.^16^ Cardiometabolic index (CMI) was calculated using the product of two ratios (waist/height x triglycerides/HDL). ^16,27^ Additional details on medications can be found in the Supplementary Material.

### 2.5 Imaging Biomarkers

#### 2.5.1 MRI Acquisition and Processing

Participants were scanned on a research-dedicated 3-Tesla Siemens Skyra magnetic resonance imaging (MRI; 32-channel head coil) scanner. Detailed image acquisition parameters have been previously published.^20,21,28^ T1, T2 FLAIR, and DTI/NODDI scans were acquired. T1 image processing included SPM12 (www.fil.ion.ucl.ac.uk/spm) CAT12 normalization and tissue segmentation, as well as regional volume and cortical thickness and total intracranial volume (ICV) estimation using FreeSurfer v7.2 (https://surfer.nmr.mgh.harvard.edu). FreeSurfer bilateral and unilateral (left and right) total cortical gray matter (GM) volume, bilateral hippocampal volume, and temporal lobe cortical thickness (bilateral entorhinal, inferior/middle temporal, fusiform) were calculated. WMH volume (WMH; lesions of presumed ischemic origin) were segmented by the lesion growth algorithm (LGA) implemented in the LST toolbox v2.0.15, running in SPM12 using FLAIR and T1 images.^29^ A log transformation was applied to WMH to account for its skewed distribution ^30,31^. DTI and NODDI processing details have been described previously.^20,21^ Briefly, NODDI free water (FW) was averaged in global supratentorial GM, global supratentorial WM, and bilateral hippocampal GM; DTI fractional anisotropy (FA) and mean diffusivity (MD) were averaged in global supratentorial WM. The Johns Hopkins University (JHU) DTI atlas^32^ and AAL atlas^33,34^ were overlaid on template-space parameter images (FW, FA or MD) that were tissue-specific (WM or GM) to extract mean signal across all supratentorial WM tracts and supratentorial GM regions of interest (ROIs), respectively. The same process was used to calculate mean global GM and WM cerebral blood flow (CBF) from Arterial Spin labelling scans using a multiphase pseudo-continuous arterial spin labeling (MP-PCASL) sequence as published in greater detail elsewhere.^20^ MRI-derived vascular biomarkers included WMH volume, global WM DTI metrics (FA, MD), NODDI free water in GM and WM, and global GM and WM CBF. MRI-derived vascular biomarkers selected to represent the V component of the ATN(V) framework, capturing complementary features of small-vessel disease pathology, included WMH volume, global WM DTI metrics (FA, MD), NODDI free water in GM and WM, and global GM and WM CBF.

#### 2.5.2 A*β*-PET Imaging

As previously described^21,35^ fibrillar A*β* brain deposition on PET was assessed with [^11^C]-Pittsburgh compound B (PiB).^36^ Participants were injected with an intravenous bolus of ∼10mCi (approximately 370 MBq) PiB. Following injection, a computed tomography (CT) scan was acquired for attenuation correction. The participant was then scanned from 40–70 minutes (6×5-min frames) post-injection on a 64-slice GE Discovery MI DR PET/CT scanner. Each participant’s CT image was coregistered to their structural MRI, and PET frames were coregistered to MRI space using the affine matrix from the CT-MRI coregistration. Aβ deposition was quantified using a voxelwise standardized uptake volume ratio (SUVr), calculated as the PiB SUVr (40-70 min, cerebellar gray matter as the reference region) signal averaged from a cortical meta-ROI sensitive to the early pathogenesis of AD relative to the uptake in the cerebellum, using FreeSurfer-segmented (v7.2; https://surfer.nmr.mgh.harvard.edu) regions.^35,37^ Centiloid (CL) values were also calculated to facilitate harmonization across ligands and centers.^38^ CL analysis was conducted in PMOD v4.1 (PMOD LLC Technologies, Switzerland). The 3D T1-weighted MRI and an aligned average of the 50-70min PET scan frames were input into the PMOD PNEURO Stepwise Maximum Probability Atlas workflow. Briefly, MRIs were normalized to the MNI-space template and segmented. Coregistered PETs were normalized to MNI space using MRI parameters. A PMOD SUVr was calculated using a standard MNI-space Centiloid ROI with the whole cerebellum as a reference region. Centiloid scores were then calculated using Klunk et al. equation 1.3b. CL =100 (PiB-SUVr-1.009)/1.067.^38^ The Centiloid processing method and atlas template was validated using the Global Alzheimer’s Association Interactive Network (GAAIN) data set.

#### 2.5.3 Tau-PET Imaging

Tau PET with [18F]Flortaucipir (FTP) was used for assessing tau neurofibrillary tangle deposition.^39,40^ Participants were injected with an IV bolus of ∼10mCi (370 MBq) (+/-10%) [18F]FTP over 5-10s, followed by 75-min uptake. A computed tomography (CT) scan was done prior to PET for attenuation correction. Emission images were acquired continuously for 75-105 min post-injection (6×5-min frames) on a 64-slice GE Discovery MI DR PET/CT scanner. Each participant’s PET frames were motion-corrected and coregistered to MRI space using rigid matching (PMOD, v.4.1). A voxelwise 80–100-minute standardized uptake volume ratio (SUVr) image was then generated with inferior cerebellar gray matter as the reference region. Temporal meta-region of interest (meta-ROI) tau deposition was calculated using FreeSurfer-segmented regions (entorhinal cortex, amygdala, inferior/middle temporal gyri, fusiform gyrus, and parahippocampal gyrus, approximating Braak I-IV), as described previously.^41–43^

#### 2.5.4 Biomarker Classification

In the current analyses, biomarker classification (e.g., ATN) was based exclusively on neuroimaging data. Amyloid (PET; [A]), tau (PET; [T]), and neurodegeneration (MRI; [N]) positivity (A+/T+/N+) was defined using a combination of visual reads and both well-defined and sample-specific a priori thresholds.^44–46^ Visual reads were conducted by (MMR, SNL, and JRB) in accordance with the prescribing information for Tauvid (FRP; Eli Lilly and Company, 2020). Sample-derived thresholds were validated using a combination of gaussian-mixture modeling and receiver operating characteristic curves using published methods.^16,21,47^ For primary analyses assessing ARTS within the ATN classification framework, amyloid-PET positivity was determined via visual read (corresponding to an approximately global SUVr ≥ 1.21^16^). Tau-PET positivity was defined by a meta-temporal SUVr ≥ 1.21).^48^ Our primary index of MRI-based neurodegeneration positivity (N+) was defined as hippocampal volume, adjusted for intracranial volume, less than 45% (e.g., HCV <= .454) of total brain volume.^49,50^ We also explored defining N+ based on a combination of ICV-adjusted global and hippocampal gray matter volumes and cortical thickness.^49,50^ There was high agreement between methods for classifying N+ (91% of cases were N+ across approaches) and mean ARTS scores were nearly identical across the two classification schemes (Mean ARTS: HCV-only N+: = –0.309, N- = .393; Combined N+ = −.294, N- = −.400), thus this method is not discussed further. Plasma AD/ADRD biomarkers, described further below were included in confirmatory and exploratory analyses.

### 2.6 Plasma Biomarkers

#### Plasma biomarkers were measured in units pg/mL

A subset of the sample with MRI and amyloid and tau PET had plasma biomarker data (p-tau217, *n*=184; A*β*42/A*β*40, NfL and GFAP, *n*=166). Plasma AD/ADRD biomarkers were collected from participants after a minimum 8-hour (water only) fast. Blood was processed within 30 minutes of collection as described previously.^16,21^ Batch shipments were sent to the National Centralized Repository for Alzheimer’s Disease and Related Dementias (NCRAD) Biomarker Assay Laboratory for analysis. Plasma A*β*42/40, NfL and GFAP were assessed using the Quanterix Simoa Neurology 4-Plex E and p-tau181 v2 Advantage Kits on a Quanterix Simoa HD-X.^21^ Plasma p-tau217 samples, collected from 2017-2023, were processed in duplicate using ALZpath Simoa p-tau 217 v2 assay kits on a Quanterix HD-X at Neurocode (Bellingham WA).^16^ P-tau217 analyses were conducted on 1st thawed samples with the same platform and assay lot number across samples. Duplicates with CVs>20% (maximum upper limit)^51–53^ or missing one value were repeated. Kit QC controls were run with each plate. The coefficient of variation (CV; mean = 4.47; standard deviation (SD) = 3.64) for p-tau217 was well within previously established reference limits and similar to other cohorts, as recently published.^54^ All plasma biomarker concentrations were quantified in pg/mL, consistent with standard reporting for Simoa-based assays. P-tau217 served as our primary plasma AD-specific biomarker in the current study.^16,54,55^ We also evaluated the p-tau217/A*β*42 ratio.^56^

### 2.7 ARTS Score Calculation

ARTS is a supervised machine learning based classifier of brain arteriolosclerosis. ARTS was trained on ex-vivo neuropathological data with demonstrated good performance predicting in-vivo brain arteriolosclerosis.^12,13,57^ Raw T1, FLAIR, and TOPUP Eddy Current Corrected DTI images were input into the ARTS program. After standard preprocessing, global WMH burden and white-matter tract-based fractional anisotropy was calculated and entered as predictors in to the ARTS classifier along with participant age at time of scan and self-reported sex (see https://markvcid.partners.org/markvcid1-protocols-resources; https://markvcid.partners.org/sites/default/files/markvcid2/protocols/ARTS_protocol_v7.13.23.pdf).^12^ ARTS outputs a single score per scan session; higher ARTS scores indicate a higher likelihood of arteriolosclerosis.

### 2.8 Statistical Analysis

WF ADRC participants (n=238) who received cognitive diagnoses at adjudication (see Cognitive Status) by June 1st, 2025, and had an MRI and both amyloid and tau PET within one year (see Figure S1 for greater details) were selected for the ARTS ATN sub-study. Participant demographics were compared across cognitive status groups and by ATN classification using chi-square tests and one-way analysis of variance. Associations between biomarkers were evaluated using Spearman’s rank correlation, appropriate for non-parametric and potentially non-linear relationships consistent with our prior work.^16^ Multivariable general linear models (GLM) examined relationships between neuroimaging and plasma variables with ARTS in unadjusted (Model 1; see Table 1), and adjusted models controlling for age, sex, race, education, *APOE*-ε4 carrier status, and days between PET acquisition or blood-sample collection and MRI scans (Model 2). Plasma biomarker models additionally controlled for body mass index (BMI) and kidney function (Estimated Glomerular Filtration Rate; eGFR) as these factors can influence plasma biomarker levels due to physiological reasons (i.e. renal function or blood volume).^21,55,58,59^ Gamma regression models were also fit when comparing ARTS and PET biomarkers given the severity of skewness in amyloid-PET and tau-PET estimates. In post-hoc analyses, we examined if associations between ARTS and AD/ADRD biomarkers were modified by cognitive status, sex, race, and *APOE*-ε4 status. Significance level was set at an alpha of α=0.05. Multiple comparisons corrections were performed across all statistical tests assessing association between ARTS and MRI, PET, and plasma AD/ADRD biomarkers using the Benjamini-Hochberg false-discovery rate (*q*).^60^ All statistical tests were conducted in R (RStudio Team, 2020).

**Table 1.** Participant Characteristics.

|  | N | OVERALL<br>N=238<sup>a</sup> | CU<br>n=132<sup>a</sup> | MCI<br>n=80<sup>a</sup> | DEM<br>n=26<sup>a</sup> | p<sup>b</sup> |
| --- | --- | --- | --- | --- | --- | --- |
| AGE (MRI) | 238 | 71 (8) | 71 (8) | 72 (7) | 70 (8) | .200 |
| SEX:FEMALE | 238 | 150 (63%) | 95 (72%) | 41 (51%) | 14 (54%) | .006 |
| RACE | 238 |  |  |  |  | .200 |
| White |  | 185 (78%) | 99 (75%) | 62 (78%) | 24 (92%) |  |
| AA/BLACK |  | 50 (21%) | 32 (24%) | 16 (20%) | 2 (7.7%) |  |
| AI/AN |  | 1 (0.4%) | 0 (0%) | 1 (1.3%) | 0 (0%) |  |
| NH/PI |  | 1 (0.4%) | 0 (0%) | 1 (1.3%) | 0 (0%) |  |
| Asian |  | 1 (0.4%) | 1 (0.8%) | 0 (0%) | 0 (0%) |  |
| EDUCATION | 238 | 15.91 (2.57) | 16.08 (2.40) | 15.61 (2.70) | 15.96 (3.03) | .500 |
| mPACC5 | 236 | -0.59 (1.13) | 0.06 (0.67) | -1.07 (0.88) | -2.51 (0.77) | <.001 |
| SMOKER | 238 | 76 (32%) | 38 (29%) | 27 (34%) | 11 (42%) | .400 |
| APOE-ε4+ | 229 | 74 (32%) | 34 (27%) | 29 (38%) | 11 (44%) | .110 |
| ARTS | 238 | -0.37 (0.17) | -0.38 (0.15) | -0.35 (0.18) | -0.33 (0.16) | .200 |
| SYSBP | 238 | 135 (21) | 133 (22) | 138 (20) | 137 (20) | .150 |
| HTN | 238 | 159 (67%) | 79 (60%) | 63 (79%) | 17 (65%) | .016 |
| LDL | 251 | 102.98 (31.28) | 103.32 (31.40) | 99.99 (31.68) | 110.69 (28.99) | .300 |
| HDL | 251 | 63.73 (19.69) | 63.94 (19.98) | 62.77 (17.78) | 65.65 (24.11) | >.90 |
| OGTT (BSL) | 255 | 97.24 (16.62) | 96.56 (16.16) | 99.71 (18.38) | 93.22 (12.06) | .300 |
| IGT | 238 | 141 (59%) | 77 (58%) | 52 (65%) | 12 (46%) | .400 |
| BMI | 238 | 27.8 (6.4) | 28.8 (6.4) | 26.9 (6.9) | 25.5 (3.7) | .009 |
| eGFR | 230 | 78 (15) | 77 (14) | 78 (16) | 81 (16) | .200 |
| HX TBI | 238 | 6 (2.5%) | 3 (2.3%) | 2 (2.5%) | 1 (3.8%) | .800 |
| HX SLEEP APNEA | 238 | 67 (28%) | 34 (26%) | 27 (34%) | 6 (23%) | .400 |
| PET IMAGING |  |  |  |  |  |  |
| Aβ+ (Visual Read) | 238 | 102 (43%) | 39 (30%) | 43 (54%) | 20 (77%) | <.001 |
| Aβ+ (CL > 24) | 233 | 85 (36%) | 30 (23%) | 37 (47%) | 18 (69%) | <.001 |
| Aβ-PET (SUVR) | 234 | 1.40 (0.45) | 1.26 (0.34) | 1.50 (0.48) | 1.79 (0.57) | <.001 |
| Aβ-PET (Centiloids) | 233 | 28 (44) | 14 (31) | 38 (47) | 67 (56) | <.001 |
| Aβ-PET MRI INT | 238 | 24 (222) | 19 (235) | 13 (231) | 79 (62) | .900 |
| TAU-PET+ (SUVR) | 238 | 97 (41%) | 33 (25%) | 49 (61%) | 15 (58%) | <.001 |
| TAU PET (SUVR) | 238 | 1.27 (0.28) | 1.17 (0.08) | 1.31 (0.22) | 1.63 (0.55) | <.001 |
| TAU-PET MRI INT | 238 | 86 (107) | 85 (117) | 81 (96) | 102 (86) | .900 |
| NEUROIMAGING |  |  |  |  |  |  |
| GMV | 238 | 0.68 (0.04) | 0.69 (0.04) | 0.67 (0.04) | 0.65 (0.04) | <.001 |
| HCV | 238 | 0.48 (0.08) | 0.51 (0.07) | 0.46 (0.07) | 0.40 (0.08) | <.001 |
| CORTTHICK | 238 | 2.75 (0.15) | 2.78 (0.13) | 2.75 (0.15) | 2.58 (0.18) | <.001 |
| WMH (log/ICV) | 238 | -1.69 (1.26) | -1.91 (1.23) | -1.45 (1.34) | -1.42 (0.94) | .025 |
| DTI FA | 238 | 0.512 (0.020) | 0.513 (0.020) | 0.510 (0.021) | 0.509 (0.019) | .500 |
| DTI MD | 238 | 0.82 (0.05) | 0.80 (0.04) | 0.82 (0.05) | 0.84 (0.04) | <.001 |
| NODDI FW | 237 | 0.173 (0.022) | 0.170 (0.022) | 0.173 (0.020) | 0.187 (0.024) | .004 |
| ASL WM CBF | 234 | 17.7 (3.3) | 18.2 (3.5) | 17.1 (2.9) | 16.8 (3.0) | .039 |
| ASL GM CBF | 234 | 36 (8) | 37 (8) | 34 (7) | 31 (6) | <.001 |
| PLASMA |  |  |  |  |  |  |
| p-tau217 | 184 | 0.45 (0.36) | 0.36 (0.26) | 0.57 (0.39) | 0.66 (0.58) | .003 |
| p-tau217/Aβ42 | 158 | 0.08 (0.07) | 0.06 (0.05) | 0.11 (0.09) | 0.12 (0.11) | .008 |
| Aβ42/40 | 166 | 0.053 (0.010) | 0.053 (0.010) | 0.053 (0.010) | 0.048 (0.008) | .018 |
| NfL | 166 | 16 (7) | 14 (6) | 18 (9) | 21 (8) | <.001 |
| GFAP | 166 | 136 (68) | 127 (63) | 138 (60) | 179 (95) | .038 |

*Abbreviations:* CU = Cognitively Unimpaired; MCI = Mild Cognitive Impairment; DEM = Dementia; ARTS = Arteriosclerosis Score; SYSBP = Systolic Blood Pressure; HTN = Hypertension; LDL = Low-Density Lipoprotein; HDL = High-Density Lipoprotein; OGTT = Oral Glucose Tolerance Test; IGT = Impaired Glucose Tolerance; BMI = Body Mass Index; eGFR = Estimated Glomerular Filtration Rate; HX TBI = History of Traumatic Brain Injury; Aβ = Amyloid Beta; SUVR = Standardized Uptake Volume Ratio; CL = Centiloids; GMV = Total Gray Matter Brain Volume Adjusted for Intracranial Volume; HCV = Hippocampal Volume Adjusted for Intracranial Volume; CORTTHICK = Cortical Thickness; WMH = Log-Transformed White Matter Hyperintensity Volume Adjusted for Intracranial Volume; DTI FA = Diffusion Tensor Imaging

## 3 RESULTS

### 3.1 Participant Characteristics

A total of 238 participants completed brain MRI and both PiB (amyloid) PET (43% A+) and FTP (tau) PET (41% T+) within one year of MRI, and all underwent clinical and cognitive evaluation (Table 1). The mean age of participants was approximately 71 years, 21% were Black, 63% were Female, and 32% were *APOE*-ε4 carriers (*APOE*-ε4+). Regarding cardiometabolic health, 67% had HTN, and 32% with a history of smoking. Demographics were comparable to and representative of the larger WF ADRC cohort (see Table S1). Participants were adjudicated by NIA-AA criteria as having normal cognition (*n*=132 [55%]), mild cognitive impairment (MCI; *n*=80 [34%]), or dementia (DEM; n=26 [11%]). CU had a higher proportion of females compared to MCI and DEM groups. MCI and DEM participants had higher blood pressure compared to CU, while MCI had the highest proportion of participants with HTN (71%; Table 1). Neuroimaging and plasma biomarkers differed by cognitive status as previously reported in the larger WF ADRC cohort ^21,61^ and were altered in the impaired groups (CU < MCI < DEM) for neuroimaging measures, except for DTI FA (*p*>.05), and each of the plasma biomarkers assessed including p-tau217, Aβ42/40, NfL, and GFAP (*all p*<.05).

### 3.2 ARTS Scores

#### 3.2.1 General Descriptives

ARTS scores ranged from −.75 to .10 (mean ± SD = −0.37±.17), suggesting a lower overall likelihood of arteriolosclerosis in our cohort compared to the reference samples, which were slightly older than our ADRC cohort (see Methods and Discussion). ARTS scores were higher on average in females, participants with HTN, and participants with a history of smoking (all *p*<.05; see Table 1 and Figure S2). ARTS scores did not differ by cognitive status, race, *APOE*-ε4 carriership, impaired glucose status, or hypertensive medication use (independent of hypertensive status), or a history of TBI or sleep apnea (*all p*>.05). Consistent with ARTS as an aging vascular biomarker, ARTS scores were strongly positively associated with age (*rho*=0.62, *p*<.001; Figure S3). ARTS was also negatively correlated with mPACC5 scores in unadjusted models (*rho*=.17, *p*=.011). Finally, ARTS scores were not associated with lipids indexed by low-and high-density lipoprotein cholesterol, nor with cardiometabolic status (e.g., CMI; all *p*>.05). Raw unadjusted Spearman correlation matrices are presented in Figure S4.

### 3.3 Biomarkers

#### 3.3.1 Associations of ARTS with Amyloid and Tau PET deposition

Higher ARTS score was significantly associated with greater levels of amyloid deposition and greater tau PET deposition in unadjusted models (Table 2, Model 1; Figure 1; also see Figure S5 [panel a] and S6), and in models adjusted by sex, race, education, *APOE*-ε4 carriership, and the interval in days between PET and MRI (Model 2). Results were nearly identical using gamma regression (see Table S2). Associations of ARTS with PET positivity of A+ and T+ are presented in the Supplemental Material (also see Table S3).

**Figure 1.**
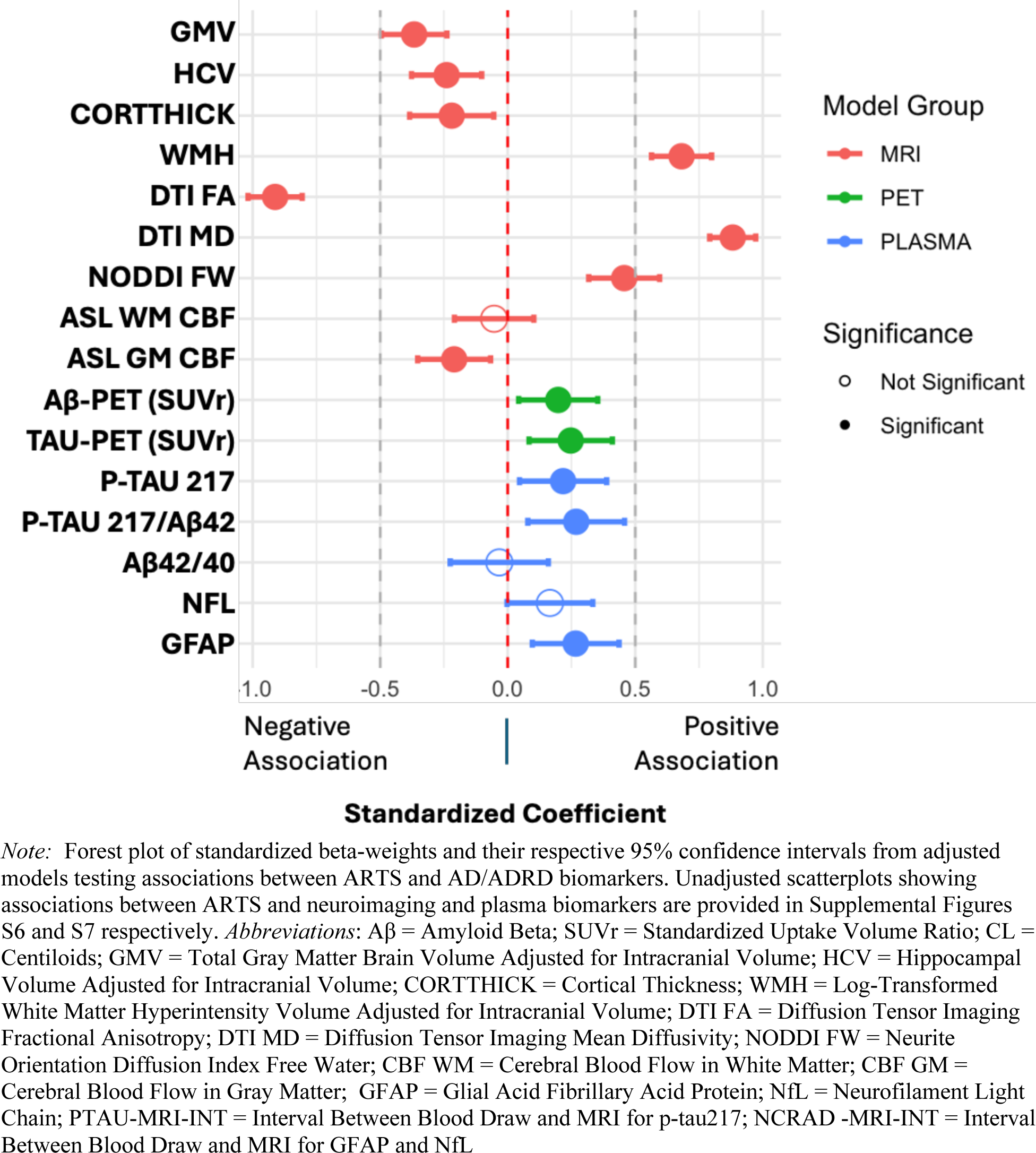
Forest plot of adjusted models evaluating associations between ARTS and AD/ADRD biomarkers.

**Table 2:** Association of ARTS with Amyloid and Tau PET deposition.

|  | Model 1 (Unadjusted) |  |  |  |  |  | Model 2 (Adjusted) |  |  |  |  |  |
| --- | --- | --- | --- | --- | --- | --- | --- | --- | --- | --- | --- | --- |
|  | *n* | R2 | *β* | *SE* | *p* | *q* | *n* | R2 | *β* | *SE* | *p* | *q* |
| **Aβ (SUVR)** | 234 | .07 | 0.70 | 0.17 | <.001 | <.001 | 225 | .22 | 0.54 | 0.22 | .013 | .017 |
| **Aβ (CL)** | 233 | .06 | 63.26 | 16.73 | <.001 | <.001 | 225 | .20 | 56.15 | 21.16 | .009 | .011 |
| **TAU (SUVR)** | 238 | .02 | 0.24 | 0.10 | .020 | .024 | 229 | .10 | 0.41 | 0.14 | .003 | .005 |

*Note:* All models were fit using robust standard errors. Model 1 was unadjusted. Model 2 was adjusted for age, sex, race, education, APOE, and the interval between PET and MRI scan acquisition. Covariates significantly contributing to model performance are summarized in Table S4. Abbreviations: R2 = Coefficient of Determination (% variance explained); SE = Standard Error; q = FDR-corrected p-values; SUVR = Standard-Uptake Value Ratio; CL = Centiloids

#### 3.3.2 Associations of ARTS with MRI measures

Higher ARTS was significantly associated with poorer overall brain health including reduced gray matter volume, DTI fractional anisotropy, and cerebral blood flow in gray matter, and higher white matter hyperintensity volume and NODDI free water across all models assessed (Table 3; Figure 1). ARTS was not significantly associated with cerebral blood flow in white matter before or after adjustment (all *p*>.05; Table 3; also see Figure S5 [panel a] and S6).

**Table 3:** Associations between ARTS and MRI measures.

| A. Structural | Model 1 (Unadjusted) |  |  |  |  |  | Model 2 (Adjusted) |  |  |  |  |  |
| --- | --- | --- | --- | --- | --- | --- | --- | --- | --- | --- | --- | --- |
|  | *n* | R2 | *β* | *SE* | *p* | *q* | *n* | R2 | *β* | *SE* | *p* | *q* |
| **GMV** | 238 | .21 | -0.12 | 0.02 | <.001 | <.001 | 229 | .43 | -0.10 | 0.02 | <.001 | <.001 |
| **HCV** | 238 | .10 | -0.14 | 0.03 | <.001 | <.001 | 229 | .36 | -0.11 | 0.03 | .001 | .001 |
| **CORTTHICK** | 238 | .05 | -0.20 | 0.06 | .001 | .002 | 229 | .07 | -0.21 | 0.01 | .008 | .011 |
| **WMH** | 238 | .48 | 5.24 | 0.36 | <.001 | <.001 | 229 | .52 | 4.96 | 0.44 | <.001 | <.001 |
| **DTI FA** | 238 | .45 | -0.08 | 0.01 | <.001 | <.001 | 229 | .61 | -0.11 | 0.01 | <.001 | <.001 |
| **DTI MD** | 238 | .63 | 0.22 | 0.01 | <.001 | <.001 | 229 | .72 | 0.25 | 0.01 | <.001 | <.001 |
| **NODDI FW** | 237 | .26 | 0.07 | 0.01 | <.001 | <.001 | 228 | .34 | 0.06 | 0.01 | <.001 | <.001 |
| B. Functional | Model 1 (Unadjusted) |  |  |  |  |  | Model 2 (Adjusted) |  |  |  |  |  |
|  | *n* | R2 | *β* | *SE* | *p* | *q* | *n* | R2 | *β* | *SE* | *p* | *q* |
| **CBF WM** | 234 | 0.00 | -0.70 | 1.32 | .598 | .616 | 225 | 0.19 | -1.14 | 1.60 | .475 | .505 |
| **CBF GM** | 234 | 0.03 | -8.38 | 3.04 | .006 | .009 | 225 | 0.31 | -9.81 | 3.50 | .005 | .008 |

*Note:* Associations between ARTS and MRI measures before and after adjustment. ARTS was most strongly associated with WMH and DTI metrics as expected (see Methods; also see Discussion for caveats). Covariates significantly contributing to model performance are summarized in Table S4. Abbreviations: Model 1 = Unadjusted; Model 2 = Adjusted for age, sex, race, education, APOE; R2 = Coefficient of Determination (% variance explained); SE = Standard Error; q = FDR-corrected p-values; CORTTHICK = Temporal Meta-

#### 3.3.3 Associations of ARTS with plasma AD/ADRD biomarkers

Higher ARTS scores were significantly associated with elevated plasma biomarker levels in unadjusted models (Model 1; Table 4, Figure 1) and models adjusted for age, sex, race, education, APOE genotype, BMI, eGFR, and the time interval between blood draw and MRI acquisition (Model 2; Table 4, Figure 1; also see Figure S5 [panel b] and S7).

**Table 4:** Associations between ARTS and plasma AD/ADRD biomarker levels.

|  | Model 1 (Unadjusted) |  |  |  |  |  | Model 2 (Adjusted) |  |  |  |  |  |
| --- | --- | --- | --- | --- | --- | --- | --- | --- | --- | --- | --- | --- |
| | *n* | R2 | $\beta$ | SE | *p* | *q* | *n* | R2 | $\beta$ | SE | *p* | *q* |
| **p-tau217** | 184 | .06 | 0.52 | 0.16 | **.001** | **.002** | **180** | .24 | 0.47 | 0.19 | **.015** | **.018** |
| **p-tau217/Aβ42** | 158 | .06 | .10 | .03 | **.002** | **.003** | **155** | .23 | 0.11 | 0.04 | **.007** | **.010** |
| **Aβ42/40** | 166 | .03 | -.01 | .00 | **.026** | **.030** | **160** | .15 | -0.00 | 0.01 | .672 | .672 |
| **NfL** | 166 | .10 | 14.01 | 3.22 | **<.001** | **<.001** | **160** | .36 | 6.34 | 3.37 | .062 | .068 |
| **GFAP** | 166 | .20 | 176.65 | 27.71 | **<.001** | **<.001** | **160** | .34 | 107.65 | 34.83 | **.002** | **.004** |

*Note:* All models were fit using robust standard errors. Model 1 was unadjusted. Model 2 was adjusted for age, sex, race, education, APOE, BMI, eGFR, and the interval between blood collection and MRI scan acquisition. Covariates significantly contributing to model performance are summarized in Table S4. Abbreviations: R2 = Coefficient of Determination (% variance explained); SE = Standard Error; *q* = FDR-corrected *p*-values; VISREAD = Visual Read; SUVR = Standard-Uptake Value Ratio; CL = Centiloids

### 3.4 ARTS and ATN Framework

ARTS scores were elevated in biomarker positivity groups (A+T-N-[*n*=25], A+T+N-[n=27], A+T+N+ [40]) as compared to A-T-N-(*n*=85; *all p*<.05). ARTS scores showed a small stepwise increase across ATN groups, consistent with a monotonic trend *(β*=0.043 per step, *p* < 0.001).

Pairwise contrasts confirmed that the largest difference was between biomarker-negative (A-T-N-) and fully biomarker-positive (A+T+N+) participants, with intermediate groups showing smaller or nonsignificant differences (Figure 2). ARTS scores tended to be higher on average in N+ groups, including those not in the canonical ATN grouping framework for AD (e.g., AD/ADRD; A-T+N-, A-T-N+, A-T+N+, A+T-N+; see Supplementary Table S5 and Table S6 for ARTS and cohort descriptives across both canonical and all ATN categorizations respectively).

**Figure 2.**
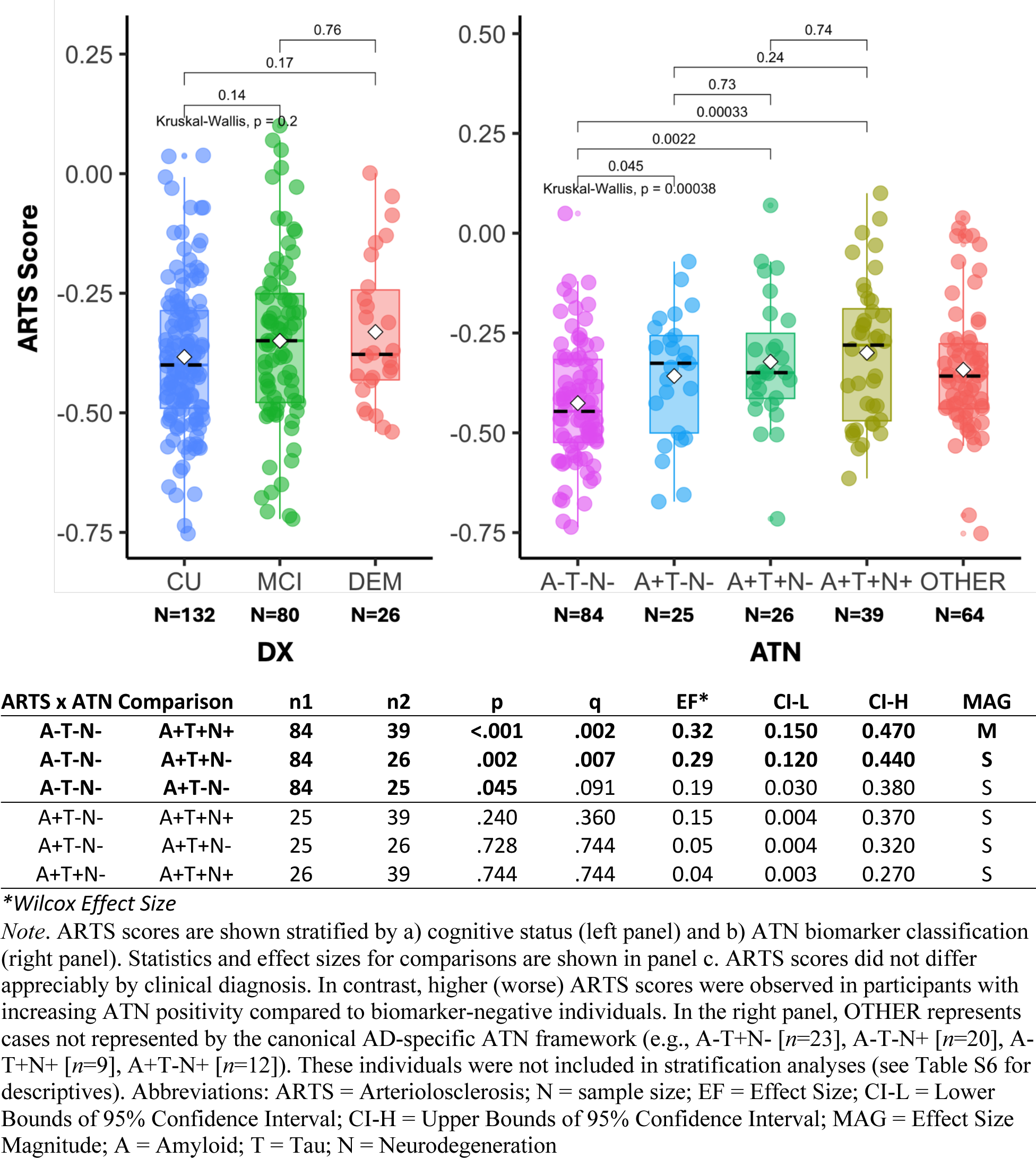
ARTS scores across diagnostic and biomarker groups.

### 3.5 Post-hoc analyses

Associations between ARTS scores and neuroimaging and plasma AD/ADRD biomarkers were largely independent of demographic, clinical, and health factors, including hypertensive status. However, in adjusted models, we observed several outcome-dependent modifications by diagnostic status, sex, and race (all interaction *p*<.05; see Figure S8). First, higher ARTS scores were more strongly associated with elevated amyloid PET levels in DEM as compared to CU and MCI participants. Second, associations between higher ARTS and lower hippocampal volume and cortical thickness estimates were stronger in female compared to male participants. Third, associations between higher ARTS scores and greater white matter hyperintensity burden were stronger in Black compared to White participants. No other interactions were observed (data not shown).

## 4 DISCUSSION

This targeted study provides novel evidence linking ARTS, a multimodal, machine learning-based index of arteriolosclerosis, to neuroimaging and plasma biomarkers of Alzheimer’s disease and related dementias (AD/ADRD) within the ATN(V) framework in a deeply phenotyped, community-based cohort. Higher ARTS scores were associated with poorer brain health, including reduced global and hippocampal brain volume, lower cortical thickness, and diminished gray matter cerebral blood flow. These structural findings are consistent with prior work demonstrating that vascular (e.g., structural) pathology contributes to neurodegeneration and cognitive decline and relates to AD pathology.^9,62–64^ In addition, higher ARTS scores were associated with putative biomarkers of V pathology (greater white matter hyperintensity burden and lower white matter microstructural integrity) as well as with plasma biomarkers of neurodegeneration (higher plasma levels of neurofilament light) and neuroinflammation (indirectly indexed by GFAP). Associations with amyloid and tau PET and with plasma p-tau217 were relatively modest and were attenuated after adjusting for cognitive status, given that these observed relationships were strongest among individuals with dementia. Finally, ARTS scores increased across the stages of the AD biomarker continuum (e.g., A+/T+/N+). By embedding ARTS within the ATN(V) framework and integrating tau/amyloid PET with plasma and perfusion metrics, this study extends prior ARTS work from structural cSVD correlates to biomarker-defined AD pathways, revealing stage-dependent and risk-modified associations.

We focused our analyses on a subset of the WF ADRC with ATN(V) biomarkers. We were particularly interested in tau PET because tau pathology is more closely linked to clinical symptoms and disease progression.^18,19,40,65–68^ Impaired clearance of amyloid-β due to vascular dysfunction may accelerate amyloid accumulation^69^ and contribute to tau progression.^70^ Evidence to date has been inconsistent regarding associations between large-vessel arteriosclerosis (e.g., arterial stiffness, pressure pulsatility) and AD/ADRD biomarkers. One study reported links with tau but not amyloid, localized to cortical regions that accumulate tau early in disease.^71^ Another found no association with tau PET burden but did observe arteriosclerosis-related amyloid deposition, modified by antihypertensive medication use, highlighting the potential importance of vascular co-pathology in AD clinical trials.^72^ Such inconsistencies likely reflect differences in cohort characteristics, the focus on large-artery disease alone, and the examination of individual etiologies in mostly unimpaired participants, where tau typically emerges later in the disease course.

This work extends prior literature with a more comprehensive view of ADRD pathology by incorporating biomarkers of arteriosclerosis and relating them to imaging and plasma measures of ATN(V). By integrating PET and plasma biomarkers, our findings extend Lamar et al. (2024), who showed ARTS predicted cognitive decline in older African Americans, and Fleischman et al. (2025), who linked ARTS to incident MCI, dementia, and stroke (see Limitations). Our findings suggest that vascular pathology, captured by ARTS (e.g., small-vessel arteriolosclerosis), may interact with tau and amyloid deposition in a stage-dependent or risk-modified manner, supporting the hypothesis that vascular dysfunction (e.g., cSVD) can exacerbate AD/ADRD pathology, particularly in individuals with midlife vascular risk factors. Because white matter hyperintensities and diffusion tensor imaging-derived fractional anisotropy are the primary imaging inputs to ARTS, strong associations with global summary metrics (e.g., WMH, DTI FA) were expected. However, it is worth noting, WMH and FA measures captured in the WF ADRC are not identical to those used in ARTS (see Methods and Limitations). Notably, ARTS scores in our cohort were lower than those reported in the initial validation by Makkinejad et al. (2021), despite ∼67% of participants having hypertension, which is a rate comparable to other evaluated cohorts and the broader WF ADRC.^12,14,57^ This discrepancy may reflect our cohort’s relatively younger age, on average, and the high prevalence of antihypertensive medication use in the WF ADRC, both of which could attenuate ARTS scores. Recently, Arfanakis and colleagues reported follow-up validation demonstrating high internal consistency and replication of ARTS scores across several cohorts independent of Makkinejad et al. (2021).^12,57^ Although these validation studies support the robustness of ARTS scores derived across different cohorts and scanner parameters, additional follow-up in the larger WF sample is needed to determine whether the lower ARTS scores in our cohort reflect scanner- or acquisition-related factors (e.g., number of diffusion directions) versus true cohort differences. Likewise, given that 94% of hypertensive individuals in our sample were taking antihypertensive medications, we were unable to distinguish controlled from uncontrolled hypertension.

Overall, ARTS scores showed small stepwise increases across ATN groups; as expected, the largest pairwise difference was observed when comparing A-T-N- to A+T+N+. ARTS scores were modestly associated with core imaging Alzheimer’s disease biomarkers and were most strongly correlated with MRI-based indicators of small-vessel disease, particularly white matter hyperintensities and fractional anisotropy (again, see Limitations), and vascular risk factors. ARTS was elevated in participants with hypertension and a history of smoking, consistent with prior work.^14,73–76^ Small-vessel injury linked to hypertension is well established to be associated with increased WMH and FA abnormalities^73–76^ and may also contribute to inflammation, as indirectly indexed by GFAP in our study. Notably, in addition to expected associations with WMH and FA, we observed relationships with NODDI Freewater and GM cerebral blood flow, suggesting that ARTS may capture broader microvascular and perfusion-related injury. These findings may raise uncertainty about the unique value of ARTS relative to established MRI and clinical measures. It remains unclear whether ARTS reflects a distinct pathophysiological signature or merely recapitulates known associations between vascular risk factors and cSVD-related neuroimaging metrics. We also did not observe associations between ARTS and lipid measures (e.g., HDL, LDL) or composite cardiometabolic indices. Lipid profiles may be more strongly linked to large-vessel atherosclerosis^77,78^ whereas associations between cSVD and cardiometabolic burden (e.g., obesity, diabetes) may be mediated by hypertension or inflammation rather than directly influencing ARTS.^79,80^ Additional work is needed to further understand the role of inflammation (I) in the context of ATN(V) and the mechanisms (e.g., endothelial dysfunction, impaired glymphatic clearance of amyloid, or neurovascular coupling) that may underlie ARTS–AD/ADRD relationships.

### Limitations and Future Directions

Our findings should be interpreted in light of limitations and technical considerations. First, the sample was restricted to participants with tau PET, reducing statistical power but enabling targeted analysis of downstream AD pathology. Although characteristics of the reduced sample were similar to the larger WF ADRC cohort, replication in larger more representative samples is needed. Although our findings suggest associations between ARTS and AD/ADRD were largely independent of demographic factors, several outcomes (e.g., sex and APOE) warrant further investigation. Likewise, given the prevalence of hypertensive medication use we were not able to assess the impact of controlled vs. uncontrolled hypertension. Second, observed associations between ARTS and amyloid PET and tau PET, and plasma p-tau217 were relatively modest in this subset of participants. Further validation of novel relationships between ARTS and AD biomarkers and consideration of cases that do not fall into the typical ATN schema are needed. Third, the cross-sectional nature of this study precludes causal inference. Longitudinal studies are required to determine whether ARTS predicts tau accumulation, amyloid progression, or cognitive decline. Fourth, ARTS is an indirect and surrogate biomarker of arteriolosclerosis derived from MRI features (WMH and FA)^12,57^ and may not capture the full spectrum of cSVD. Future work should examine its relationship to other cerebrovascular markers such as cerebral microbleeds, lacunes, and enlarged perivascular spaces.^81–84^ Moreover, vascular and metabolic comorbidities, including hypertension, diabetes, obesity, and renal dysfunction, are common in aging populations and may interact with genetic and demographic risk factors to confer greater risk of AD/ADRD.^85–88^ Advanced analytic approaches that integrate distinct imaging and clinical features could help disentangle how arteriolosclerosis specifically contributes to cognitive decline and dementia risk. Finally, peripheral and cerebral measures of arteriosclerosis can diverge, as compensatory mechanisms in the brain may partially protect cerebral vessels from systemic arterial stiffening.^89^ This distinction underscores the importance of examining both large- and small-vessel disease, which likely act through complementary but distinct pathways. In future work, we plan to directly test how microvascular injury interacts with systemic vascular changes to impact AD/ADRD pathology.

## Conclusion

To our knowledge, this is the first study to integrate the MRI-derived ARTS score with established neuroimaging and plasma biomarkers of AD/ADRD in the ATN(V) framework. Higher ARTS scores were consistently associated with poorer brain health, including reduced volumes, lower cortical thickness, diminished gray matter cerebral blood flow, and elevated plasma markers of neuroaxonal injury and neuroinflammation. Novel associations with tau PET extend prior work and situate vascular injury alongside amyloid, tau, and neurodegeneration in the ATN(V) continuum. These findings highlight the importance of addressing vascular health in midlife and support ARTS as a non-invasive, scalable biomarker of small-vessel disease. More broadly, they reinforce the hypothesis that arteriolosclerosis contributes to AD/ADRD pathology through converging vascular and neurodegenerative pathways. Future studies should determine whether modifying vascular risk alters ARTS trajectories, downstream neurodegenerative processes, and cognitive outcomes, and should test whether ARTS diverges from large-vessel atherosclerosis, commonly influenced by lipid metabolism, by capturing distinct small-vessel mechanisms such as impaired clearance, neurovascular coupling deficits, and neuroinflammation.

## Data Availability

All data produced in the present study are available upon reasonable request to the authors

## Acknowledgements

This work was supported by the Wake Forest University School of Medicine’s Alzheimer’s Disease Research Center (P30AG049638, P30AG072947, R01AG054069, and R01AG058969, which is funded by the National Institute on Aging (NIA). Additional support was provided by the Department of Gerontology and Geriatric Medicine and Center for Healthy Aging and Alzheimer’s Prevention, Wake Forest School of Medicine. This study would not have been possible without the commitment and support of our valued WF ADRC staff and study participants.

## Contributions

All authors made substantial contributions and contributed to the final draft. MDR, TMH, and SNL contributed to the conception and design. MDR performed data analyses. All authors approved the version to be published and agree to be accountable for all aspects of the work in ensuring that questions related to the accuracy or integrity of any part of the work are appropriately investigated and resolved.

## Ethics approval and consent to participate

The Wake Forest Institutional Review Board approved all activities as described; written informed consent was obtained for all participants and/or their legally authorized representatives.

## Consent for publication

Written informed consent was obtained for all participants and/or their legally authorized representatives.

## References

1. Gottesman RF, Schneider ALC, Zhou Y, et al. Association between midlife vascular risk factors and estimated brain amyloid deposition. JAMA. 2017;317(14):1443. doi:10.1001/JAMA.2017.3090

2. Sweeney MD, Montagne A, Sagare AP, et al. Vascular dysfunction—The disregarded partner of Alzheimer’s disease. Alzheimer’s and Dementia.Elsevier Inc. 2019;15(1):158–167. doi:10.1016/j.jalz.2018.07.222

3. Kapasi A, DeCarli C, Schneider JA. Impact of Multiple Pathologies on the Threshold for Clinically Overt Dementia. Acta Neuropathol. 2017;134(2):171. doi:10.1007/S00401-017-1717-7

4. Kapasi A, Leurgans SE, Arvanitakis Z, Barnes LL, Bennett DA, Schneider JA. β-amyloid and tau tangle pathology modifies the association between small vessel disease and cortical microinfarcts. Stroke. 2021;52(3):1012. doi:10.1161/STROKEAHA.120.031073

5. Burke GL, Hughes TM. Arterial Changes Connecting Hypertension to Alzheimer’s Disease and Related Dementias. JACC Cardiovasc Imaging. 2021;14(1):186–188. doi:10.1016/J.JCMG.2020.10.018

6. Nordestgaard LT, Christoffersen M, Frikke-Schmidt R. Shared Risk Factors between Dementia and Atherosclerotic Cardiovascular Disease. International Journal of Molecular Sciences 2022, Vol 23, Page 9777. 2022;23(17):9777. doi:10.3390/IJMS23179777

7. Nichols E, Merrick R, Hay SI, et al. The prevalence, correlation, and co-occurrence of neuropathology in old age: harmonisation of 12 measures across six community-based autopsy studies of dementia. Lancet Healthy Longev. 2023;4(3):e115–e125. doi:10.1016/S2666-7568(23)00019-3

8. Duering M, Biessels GJ, Brodtmann A, et al. Neuroimaging standards for research into small vessel disease—advances since 2013. Lancet Neurol. 2023;22(7):602–618. doi:10.1016/S1474-4422(23)00131-X/ATTACHMENT/44AFD1BC-CF41-4756-8886-A019A142F63A/MMC1.PDF

9. Ighodaro ET, Abner EL, Fardo DW, et al. Risk factors and global cognitive status related to brain arteriolosclerosis in elderly individuals. Journal of Cerebral Blood Flow & Metabolism. 2016;37(1):201. doi:10.1177/0271678X15621574

10. Kryscio RJ, Abner EL, Nelson PT, et al. The Effect of Vascular Neuropathology on Late-life Cognition: Results from the SMART Project. J Prev Alzheimers Dis. 2016;3(2):85. doi:10.14283/JPAD.2016.95

11. Arvanitakis Z, Capuano AW, Leurgans SE, Bennett DA, Schneider JA. Relation of Cerebral Vessel Disease to Alzheimer’s Disease Dementia and Cognitive Function in Older Persons: A Cross-sectional Study. Lancet Neurol. 2016;15(9):934. doi:10.1016/S1474-4422(16)30029-1

12. Makkinejad N, Evia AM, Tamhane AA, et al. ARTS: A novel In-vivo classifier of arteriolosclerosis for the older adult brain. Neuroimage Clin. 2021;31:102768. doi:10.1016/J.NICL.2021.102768

13. Lamar M, Arfanakis K, Evia A, et al. Changes in an in-vivo classifier of ARTerioloSclerosis (ARTS) with simultaneous change in cognition for older African Americans. Neurobiol Aging. 2024;134:21–27. doi:10.1016/J.NEUROBIOLAGING.2023.11.003

14. Fleischman DA, Arfanakis K, Leurgans SE, et al. ARTS is associated with vascular risk factors, MCI, dementia, and stroke. Alzheimer’s & Dementia. 2025;21(7):e70430. doi:10.1002/ALZ.70430

15. Jack CR, Bennett DA, Blennow K, et al. NIA-AA Research Framework: Toward a biological definition of Alzheimer’s disease. Alzheimer’s and Dementia.Elsevier Inc. 2018;14(4):535–562. doi:10.1016/j.jalz.2018.02.018

16. Rudolph MD, Sutphen CL, Register TC, et al. Evaluation of plasma p-tau217 for detecting amyloid pathology in a heterogeneous community-based cohort. Alzheimer’s & Dementia. 2025;21(7):e70426. doi:10.1002/ALZ.70426

17. Van Der Flier WM, Scheltens P. The ATN Framework—Moving Preclinical Alzheimer Disease to Clinical Relevance. JAMA Neurol. 2022;79(10):968–970. doi:10.1001/JAMANEUROL.2022.2967

18. Jack CR, Bennett DA, Blennow K, et al. A/T/N: An unbiased descriptive classification scheme for Alzheimer disease biomarkers. Neurology. 2016;87(5):539–547. doi:10.1212/WNL.0000000000002923/SUPPL_FILE/456.PDF

19. Moscoso A, Heeman F, Raghavan S, et al. Frequency and Clinical Outcomes Associated With Tau Positron Emission Tomography Positivity. JAMA. Published online June 16, 2025. doi:10.1001/JAMA.2025.7817

20. Coffin C, Suerken CK, Bateman JR, et al. Vascular and microstructural markers of cognitive pathology. Alzheimer’s & Dementia : Diagnosis, Assessment & Disease Monitoring. 2022;14(1). doi:10.1002/DAD2.12332

21. Rudolph MD, Sutphen CL, Register TC, et al. Associations among plasma, MRI, and amyloid PET biomarkers of Alzheimer’s disease and related dementias and the impact of health-related comorbidities in a community-dwelling cohort. Alzheimer’s & Dementia. 2024;20(6):4159. doi:10.1002/ALZ.13835

22. Albert MS, DeKosky ST, Dickson D, et al. The diagnosis of mild cognitive impairment due to Alzheimer’s disease: Recommendations from the National Institute on Aging-Alzheimer’s Association workgroups on diagnostic guidelines for Alzheimer’s disease. Alzheimers Dement. 2011;7(3):270. doi:10.1016/J.JALZ.2011.03.008

23. McKhann GM, Knopman DS, Chertkow H, et al. The diagnosis of dementia due to Alzheimer’s disease: Recommendations from the National Institute on Aging-Alzheimer’s Association workgroups on diagnostic guidelines for Alzheimer’s disease. Alzheimers Dement. 2011;7(3):263. doi:10.1016/J.JALZ.2011.03.005

24. Papp K V., Rentz DM, Orlovsky I, Sperling RA, Mormino EC. Optimizing the preclinical Alzheimer’s cognitive composite with semantic processing: The PACC5. Alzheimer’s and Dementia: Translational Research and Clinical Interventions. 2017;3(4):668–677. doi:10.1016/j.trci.2017.10.004

25. Donohue MC, Sperling RA, Salmon DP, et al. The preclinical Alzheimer cognitive composite: Measuring amyloid-related decline. JAMA Neurol. 2014;71(8):961–970. doi:10.1001/jamaneurol.2014.803

26. Whelton PK, Carey RM, Aronow WS, et al. 2017 ACC/AHA/AAPA/ABC/ACPM/AGS/APhA/ASH/ASPC/NMA/PCNA Guideline for the Prevention, Detection, Evaluation, and Management of High Blood Pressure in Adults: Executive Summary: A Report of the American College of Cardiology/American Heart Association Task Force on Clinical Practice Guidelines. Hypertension. 2018;71(6):1269-1324. doi:10.1161/HYP.0000000000000066

27. Wakabayashi I, Daimon T. The “cardiometabolic index” as a new marker determined by adiposity and blood lipids for discrimination of diabetes mellitus. Clinica Chimica Acta. 2015;438:274–278. doi:10.1016/J.CCA.2014.08.042

28. Hughes TM, Lockhart SN, Suerken CK, et al. Hypertensive Aspects of Cardiometabolic Disorders Are Associated with Lower Brain Microstructure, Perfusion, and Cognition. J Alzheimers Dis. 2022;90(4):1589. doi:10.3233/JAD-220646

29. Schmidt P, Gaser C, Arsic M, et al. An automated tool for detection of FLAIR-hyperintense white-matter lesions in Multiple Sclerosis. Neuroimage. 2012;59(4):3774–3783. doi:10.1016/j.neuroimage.2011.11.032

30. Raz N, Yang YQ, Rodrigue KM, Kennedy KM, Lindenberger U, Ghisletta P. White matter deterioration in 15 months: Latent growth curve models in healthy adults. Neurobiol Aging. 2012;33(2):429.e1-429.e5. doi:10.1016/j.neurobiolaging.2010.11.018

31. Raz N, Yang Y, Dahle CL, Land S. Volume of white matter hyperintensities in healthy adults: Contribution of age, vascular risk factors, and inflammation-related genetic variants. Biochimica et Biophysica Acta (BBA) - Molecular Basis of Disease. 2012;1822(3):361–369. doi:10.1016/J.BBADIS.2011.08.007

32. Oishi K, Faria A, Jiang H, et al. Atlas-based whole brain white matter analysis using large deformation diffeomorphic metric mapping: Application to normal elderly and Alzheimer’s disease participants. Neuroimage. 2009;46(2):486. doi:10.1016/J.NEUROIMAGE.2009.01.002

33. Rolls ET, Joliot M, Tzourio-Mazoyer N. Implementation of a new parcellation of the orbitofrontal cortex in the automated anatomical labeling atlas. Neuroimage. 2015;122:1–5. doi:10.1016/J.NEUROIMAGE.2015.07.075

34. Tzourio-Mazoyer N, Landeau B, Papathanassiou D, et al. Automated Anatomical Labeling of Activations in SPM Using a Macroscopic Anatomical Parcellation of the MNI MRI Single-Subject Brain. Neuroimage. 2002;15(1):273–289. doi:10.1006/NIMG.2001.0978

35. Lockhart SN, Schaich CL, Craft S, et al. Associations among vascular risk factors, neuroimaging biomarkers, and cognition: Preliminary analyses from the Multi-Ethnic Study of Atherosclerosis (MESA). Alzheimer’s & Dementia. 2022;18(4):551–560. doi:10.1002/ALZ.12429

36. Klunk WE, Engler H, Nordberg A, et al. Imaging brain amyloid in Alzheimer’s disease with Pittsburgh Compound-B. Ann Neurol. 2004;55(3):306–319. doi:10.1002/ANA.20009

37. Mormino EC, Brandel MG, Madison CM, et al. Not quite PIB-positive, not quite PIB-negative: slight PIB elevations in elderly normal control subjects are biologically relevant. Neuroimage. 2012;59(2):1152. doi:10.1016/J.NEUROIMAGE.2011.07.098

38. Klunk WE, Koeppe RA, Price JC, et al. The Centiloid Project: Standardizing quantitative amyloid plaque estimation by PET. Alzheimer’s & Dementia. 2015;11(1):1–15.e4. doi:10.1016/J.JALZ.2014.07.003

39. Lockhart SN, Schöll M, Baker SL, et al. Amyloid and Tau PET Demonstrate Region-Specific Associations in Normal Older People. Neuroimage. 2017;150:191. doi:10.1016/J.NEUROIMAGE.2017.02.051

40. Ossenkoppele R, Schonhaut DR, Schöll M, et al. Tau PET patterns mirror clinical and neuroanatomical variability in Alzheimer’s disease. Brain. 2016;139(5):1551. doi:10.1093/BRAIN/AWW027

41. Jack CR, Wiste HJ, Weigand SD, et al. Defining imaging biomarker cut-points for brain aging and Alzheimer’s disease. Alzheimers Dement. 2016;13(3):205. doi:10.1016/J.JALZ.2016.08.005

42. Jack CR, Wiste HJ, Weigand SD, et al. Age and sex specific prevalences of cerebral β-amyloidosis, tauopathy and neurodegeneration among clinically normal individuals aged 50-95 years: a cross-sectional study. Lancet Neurol. 2017;16(6):435. doi:10.1016/S1474-4422(17)30077-7

43. Leuzy A, Pascoal TA, Strandberg O, et al. A multicenter comparison of [18F]flortaucipir, [18F]RO948, and [18F]MK6240 tau PET tracers to detect a common target ROI for differential diagnosis. Eur J Nucl Med Mol Imaging. 2021;48(7):2295. doi:10.1007/S00259-021-05401-4

44. Villeneuve S, Rabinovici GD, Cohn-Sheehy BI, et al. Existing Pittsburgh Compound-B positron emission tomography thresholds are too high: statistical and pathological evaluation. Brain. 2015;138(7):2020. doi:10.1093/BRAIN/AWV112

45. Knopman DS, Lundt ES, Therneau TM, et al. Association of Initial β-Amyloid Levels With Subsequent Flortaucipir Positron Emission Tomography Changes in Persons Without Cognitive Impairment. JAMA Neurol. 2021;78(2):1. doi:10.1001/JAMANEUROL.2020.3921

46. La Joie R, Ayakta N, Seeley WW, et al. Multi-site study of the relationships between ante mortem [11C]PIB-PET Centiloid values and post mortem measures of Alzheimer’s disease neuropathology. Alzheimers Dement. 2019;15(2):205. doi:10.1016/J.JALZ.2018.09.001

47. Rudolph MD, Tanley JE, Ding J, et al. Subclinical Vascular Risk Composites and Dementia Imaging Biomarkers 17-20 years later in the Multi-Ethnic Study of Atherosclerosis (MESA). Alzheimer’s & Dementia. 2024;20(S7):e089603. doi:10.1002/ALZ.089603

48. Weigand AJ, Maass A, Eglit GL, Bondi MW. What’s the cut-point?: a systematic investigation of tau PET thresholding methods. Alzheimers Res Ther. 2022;14(1):1–17. doi:10.1186/S13195-022-00986-W/FIGURES/7

49. Schwarz CG, Gunter JL, Wiste HJ, et al. A large-scale comparison of cortical thickness and volume methods for measuring Alzheimer’s disease severity. Neuroimage Clin. 2016;11:802. doi:10.1016/J.NICL.2016.05.017

50. Allison SL, Koscik RL, Cary RP, et al. Comparison of different MRI-based morphometric estimates for defining neurodegeneration across the Alzheimer’s disease continuum. Neuroimage Clin. 2019;23:101895. doi:10.1016/J.NICL.2019.101895

51. Benedet AL, Brum WS, Hansson O, et al. The accuracy and robustness of plasma biomarker models for amyloid PET positivity. Alzheimers Res Ther. 2022;14(1):1–11. doi:10.1186/s13195-021-00942-0

52. Liu HC, Chen HH, Ho CS, et al. Investigation of the Number of Tests Required for Assaying Plasma Biomarkers Associated with Alzheimer’s Disease Using Immunomagnetic Reduction. Neurol Ther. 2021;10(2):1015. doi:10.1007/S40120-021-00280-1

53. Tworoger SS, Hankinson SE. Use of biomarkers in epidemiologic studies: Minimizing the influence of measurement error in the study design and analysis. Cancer Causes and Control. 2006;17(7):889–899. doi:10.1007/S10552-006-0035-5/METRICS

54. Ashton NJ, Wagner;, Brum S, et al. Diagnostic Accuracy of a Plasma Phosphorylated Tau 217 Immunoassay for Alzheimer Disease Pathology. JAMA Neurol. Published online January 22, 2024. doi:10.1001/JAMANEUROL.2023.5319

55. Mielke MM, Dage JL, Frank RD, et al. Performance of plasma phosphorylated tau 181 and 217 in the community. Nature Medicine 2022 28:7. 2022;28(7):1398–1405. doi:10.1038/s41591-022-01822-2

56. Wang J, Huang S, Lan G, et al. Diagnostic accuracy of plasma p-tau217/Aβ42 for Alzheimer’s disease in clinical and community cohorts. Alzheimer’s & Dementia. 2025;21(3):e70038. doi:10.1002/ALZ.70038

57. Arfanakis K, Arnold M Evia J, Leurgans SE, et al. The ARTS Marker of Arteriolosclerosis: Instrumental and Clinical Validation. Alzheimer’s & Dementia. 2025;20(Suppl 9):e094375. doi:10.1002/ALZ.094375

58. Mielke MM, Fowler NR. Alzheimer disease blood biomarkers: considerations for population-level use. Nature Reviews Neurology 2024 20:8. 2024;20(8):495-504. doi:10.1038/s41582-024-00989-1

59. Syrjanen JA, Campbell MR, Algeciras-Schimnich A, et al. Associations of amyloid and neurodegeneration plasma biomarkers with comorbidities. Alzheimer’s and Dementia. 2022;18(6):1128–1140. doi:10.1002/alz.12466

60. Benjamini Y, Hochberg Y. Controlling the false discovery rate: a practical and powerful approach to multiple testing. Journal of the Royal Statistical Society. 1995;57(1):289–300. doi:10.2307/2346101

61. Rudolph MD, Sutphen CL, Register TC, et al. Evaluation of plasma p-tau217 for detecting amyloid pathology in a diverse and heterogeneous community-based cohort. medRxiv. Published online January 1, 2025:2025.01.20.25320851. doi:10.1101/2025.01.20.25320851

62. Hughes TM, Kuller LH, Barinas-Mitchell EJM, et al. Arterial stiffness and β-amyloid progression in nondemented elderly adults. JAMA Neurol. 2014;71(5):562–568. doi:10.1001/JAMANEUROL.2014.186

63. Hughes TM, Kuller LH, Barinas-Mitchell EJM, et al. Pulse wave velocity is associated with β-amyloid deposition in the brains of very elderly adults. Neurology. 2013;81(19):1711–1718. doi:10.1212/01.WNL.0000435301.64776.37

64. Hughes TM, Wagenknecht LE, Craft S, et al. Arterial stiffness and dementia pathology: Atherosclerosis Risk in Communities (ARIC)-PET Study. Neurology. 2018;90(14):e1248. doi:10.1212/WNL.0000000000005259

65. Schöll M, Lockhart SN, Schonhaut DR, et al. PET Imaging of Tau Deposition in the Aging Human Brain. Neuron. 2016;89(5):971–982. doi:10.1016/J.NEURON.2016.01.028

66. Ossenkoppele R, Reimand J, Smith R, et al. Tau PET correlates with different Alzheimer’s disease-related features compared to CSF and plasma p-tau biomarkers. EMBO Mol Med. 2021;13(8):e14398. doi:10.15252/emmm.202114398

67. Weigand AJ, Bangen KJ, Thomas KR, et al. Is tau in the absence of amyloid on the Alzheimer’s continuum?: A study of discordant PET positivity. Brain Commun. 2020;2(1). doi:10.1093/braincomms/fcz046

68. Ioannou K, Bucci M, Tzortzakakis A, Savitcheva I, Nordberg A, Chiotis K. Tau PET positivity predicts clinically relevant cognitive decline driven by Alzheimer’s disease compared to comorbid cases; proof of concept in the ADNI study. Molecular Psychiatry 2024. Published online August 23, 2024:1–13. doi:10.1038/s41380-024-02672-9

69. Weller RO, Subash M, Preston SD, Mazanti I, Carare RO. Perivascular drainage of amyloid-beta peptides from the brain and its failure in cerebral amyloid angiopathy and Alzheimer’s disease. Brain Pathol. 2008;18(2):253–266. doi:10.1111/J.1750-3639.2008.00133.X

70. Cai Y, Du J, Li A, et al. Initial levels of β-amyloid and tau deposition have distinct effects on longitudinal tau accumulation in Alzheimer’s disease. Alzheimers Res Ther. 2023;15(1):1–14. doi:10.1186/S13195-023-01178-W/FIGURES/5

71. Cooper LL, O’Donnell A, Beiser AS, et al. Association of Aortic Stiffness and Pressure Pulsatility With Global Amyloid-β and Regional Tau Burden Among Framingham Heart Study Participants Without Dementia. JAMA Neurol. 2022;79(7):710–719. doi:10.1001/JAMANEUROL.2022.1261

72. Köbe T, Gonneaud J, Pichet Binette A, et al. Association of Vascular Risk Factors With β-Amyloid Peptide and Tau Burdens in Cognitively Unimpaired Individuals and Its Interaction With Vascular Medication Use. JAMA Netw Open. 2020;3(2):e1920780–e1920780. doi:10.1001/JAMANETWORKOPEN.2019.20780

73. Solé-Guardia G, Janssen A, Wolters R, et al. Impact of hypertension on cerebral small vessel disease: A post-mortem study of microvascular pathology from normal-appearing white matter into white matter hyperintensities. Journal of Cerebral Blood Flow & Metabolism. Published online September 1, 2025:0271678X251333256. doi:10.1177/0271678X251333256

74. Solé-Guardia G, Custers E, de Lange A, et al. Association between hypertension and neurovascular inflammation in both normal-appearing white matter and white matter hyperintensities. Acta Neuropathol Commun. 2023;11(1):2. doi:10.1186/S40478-022-01497-3

75. Nasrallah IM, Pajewski NM, Auchus AP, et al. Association of Intensive vs Standard Blood Pressure Control With Cerebral White Matter Lesions. JAMA. 2019;322(6):524–534. doi:10.1001/JAMA.2019.10551

76. Wartolowska KA, Webb AJS. Blood Pressure Determinants of Cerebral White Matter Hyperintensities and Microstructural Injury: UK Biobank Cohort Study. Hypertension. 2021;78(2):532–539. doi:10.1161/HYPERTENSIONAHA.121.17403/SUPPL_FILE/HYP_HYPE-2021-17403_SUPP1.PDF

77. Rao C, Zhu L, Yu C, et al. Association of novel lipid indices with the white matter hyperintensities in cerebral small vessel disease: a cross-sectional study. Lipids Health Dis. 2024;23(1):333. doi:10.1186/S12944-024-02318-3

78. Xie Y, Liu S, Wang X, et al. Lipids, Apolipoproteins, Lipid-Lowering Drugs, and the Risk of Cerebral Small Vessel Disease: A Mendelian Randomization Study. Journal of the American Heart Association. 2024;13(16):32409. doi:10.1161/JAHA.123.032409/SUPPL_FILE/JAH39984-SUP-0001-SUPINFO.PDF

79. Patel V, Edison P. Cardiometabolic risk factors and neurodegeneration: a review of the mechanisms underlying diabetes, obesity and hypertension in Alzheimer’s disease. J Neurol Neurosurg Psychiatry. 2024;95(6):581–589. doi:10.1136/JNNP-2023-332661

80. Ishida A, Nakanishi R, Miyagi T, et al. Association of Obesity and Metabolic Health Status with Cerebral Small-Vessel Disease in Stroke-Free Individuals. J Atheroscler Thromb. 2025;32(10):1304. doi:10.5551/JAT.65649

81. Hwang G, Abdulkadir A, Erus G, et al. Disentangling Alzheimer’s disease neurodegeneration from typical brain ageing using machine learning. Brain Commun. 2022;4(3). doi:10.1093/BRAINCOMMS/FCAC117

82. Rashid T, Liu H, Ware JB, et al. Deep learning based detection of enlarged perivascular spaces on brain MRI. Neuroimage: Reports. 2023;3(1):100162. doi:10.1016/J.YNIRP.2023.100162

83. Charisis S, Rashid T, Liu H, et al. Assessment of Risk Factors and Clinical Importance of Enlarged Perivascular Spaces by Whole-Brain Investigation in the Multi-Ethnic Study of Atherosclerosis. JAMA Netw Open. 2023;6(4):e239196. doi:10.1001/JAMANETWORKOPEN.2023.9196

84. Sudre CH, Van Wijnen K, Dubost F, et al. Where is VALDO? VAscular Lesions Detection and segmentatiOn challenge at MICCAI 2021. Med Image Anal. 2024;91:103029. doi:10.1016/J.MEDIA.2023.103029

85. Henning RJ. Obesity and obesity-induced inflammatory disease contribute to atherosclerosis: a review of the pathophysiology and treatment of obesity. Am J Cardiovasc Dis. 2021;11(4):504. Accessed May 29, 2025. https://pmc.ncbi.nlm.nih.gov/articles/PMC8449192/

86. Kadoya M, Koyama H. Sleep, Autonomic Nervous Function and Atherosclerosis. Int J Mol Sci. 2019;20(4):794. doi:10.3390/IJMS20040794

87. Full KM, Huang T, Shah NA, et al. Sleep Irregularity and Subclinical Markers of Cardiovascular Disease: The Multi-Ethnic Study of Atherosclerosis. J Am Heart Assoc. 2023;12(4):27361. doi:10.1161/JAHA.122.027361/SUPPL_FILE/JAH38121-SUP-0001-SUPINFO.PDF

88. Domínguez F, Fuster V, Fernández-Alvira JM, et al. Association of Sleep Duration and Quality With Subclinical Atherosclerosis. J Am Coll Cardiol. 2019;73(2):134–144. doi:10.1016/J.JACC.2018.10.060

89. Scheuermann BC, Parr SK, Schulze KM, et al. Associations of Cerebrovascular Regulation and Arterial Stiffness With Cerebral Small Vessel Disease: A Systematic Review and Meta-Analysis. Journal of the American Heart Association: Cardiovascular and Cerebrovascular Disease. 2023;12(23):e032616. doi:10.1161/JAHA.123.032616

90. Williams JR, Deconne TM, Pewowaruk R, et al. Total and Structural Carotid Artery Stiffness Are Associated With Cognitive Decline and Structural Brain Abnormalities Related to Alzheimer Disease and Alzheimer Disease-Related Dementias Pathology: The Multi-Ethnic Study of Atherosclerosis. Journal of the American Heart Association: Cardiovascular and Cerebrovascular Disease. 2025;14(9):e039925. doi:10.1161/JAHA.124.039925

