## Supplemental Material for "Evaluation of an in vivo biomarker of arteriolosclerosis (ARTS) and its associations with cognition and multimodal ATN(V) biomarkers in a cardiometabolic-risk enriched community cohort"

**Supplementary Materials**

**Supplemental Methods**

***Exclusion criteria.*** As previously described, exclusion criteria included a history of large vessel stroke (participants with lacunae or small vessel ischemic disease were eligible); other significant neurologic diseases; uncontrolled chronic medical or psychiatric conditions (such as advanced liver or severe kidney disease [eGFR < 30]; poorly controlled congestive heart failure, chronic obstructive pulmonary disease or sleep apnea; active cancer treatment; uncontrolled clinical depression, or psychiatric illness; current use of insulin; and history of substance abuse or heavy alcohol consumption within the previous ten years).[1–4]

***Medications***. Participants self-reported all prescription and over-the-counter medications taken and were considered to be on anti-hypertensive therapy if taking any of the following: antiadrenergic agents, angiotensin converting enzyme inhibitors, beta-blockers, calcium channel blocking agents, diuretics, vasodilators, angiotensin II inhibitors, or antihypertensive combination therapy agents. As previously described,[2,3] participants were considered on antihypertension therapy if they reported recent or current use of antiadrenergic agents, angiotensin-converting enzyme inhibitors, beta-blockers, calcium channel blocking agents, diuretics, vasodilators, angiotensin II inhibitors, or antihypertensive combination therapy agents.

***Amyloid and tau PET Imaging.*** Amyloid and tau PET measurements are evaluated both as continuous and categorical biomarkers; the marked skewness and presence of a biological threshold where deposition shifts from baseline to pathologic levels justify modeling these biomarkers as categorical outcomes or using non-parametric approaches given log or other transformations are often ineffective. Further, although it is often not recommended to dichotomize continuous measures, categorical cut-points are necessary and required in clinical setting and in clinical decision-making. Here we evaluated associations between ARTS and amyloid and tau PET burden using traditional GLMs, Poisson and GAMMA regression, and logistic regression. Overall, results were largely consistent. Supported by supplementary analyses, where ARTS was more strongly associated with amyloid and tau PET deposition (1) in the upper quartiles of the distribution for each biomarker and (2) in younger adults who self-reported taking hypertensive medications. Although these preliminary findings were exploratory, they are supported by previous literature; follow-up evaluation will be critical.

**Supplementary Results**

Linear regression models estimating relationship between ARTS and amyloid and tau-PET deposition exhibited poor fit; residuals deviated significantly from a normal distribution (Shapiro-Wilk normality test: $p < 0.05$) and there was evidence of heteroscedasticity (Breusch-Pagan test: $p = .024$). As expected, log-transformation had no impact on the distribution of amyloid or tau-PET values. The association between ARTS and amyloid and tau-PET deposition was not significant using robust and Gamma regression modeling (both $p > .05$; Table S4).

**3.3.2 Association of ARTS with Amyloid and Tau PET positivity**

Higher ARTS scores were significantly associated with an increased odds of being classified as amyloid and tau PET positive in unadjusted models (Table S5; Model 1). In models adjusted for age, sex, race, education, APOE, and the interval between PET and MRI acquisition (Table S5; Model 2), higher ARTS scores were also associated with amyloid PET positivity when using a more lenient Centiloid threshold of 12.2, corresponding with CERAD plaque scores (see Methods), and with tau PET positivity based on visual reads (*both unadjusted p*<.05). However, these associations did not survive multiple comparisons corrections.

**Supplemental Figures & Tables**

**Figure S1.** Distribution of days between amyloid and tau PET scans and closest MRI

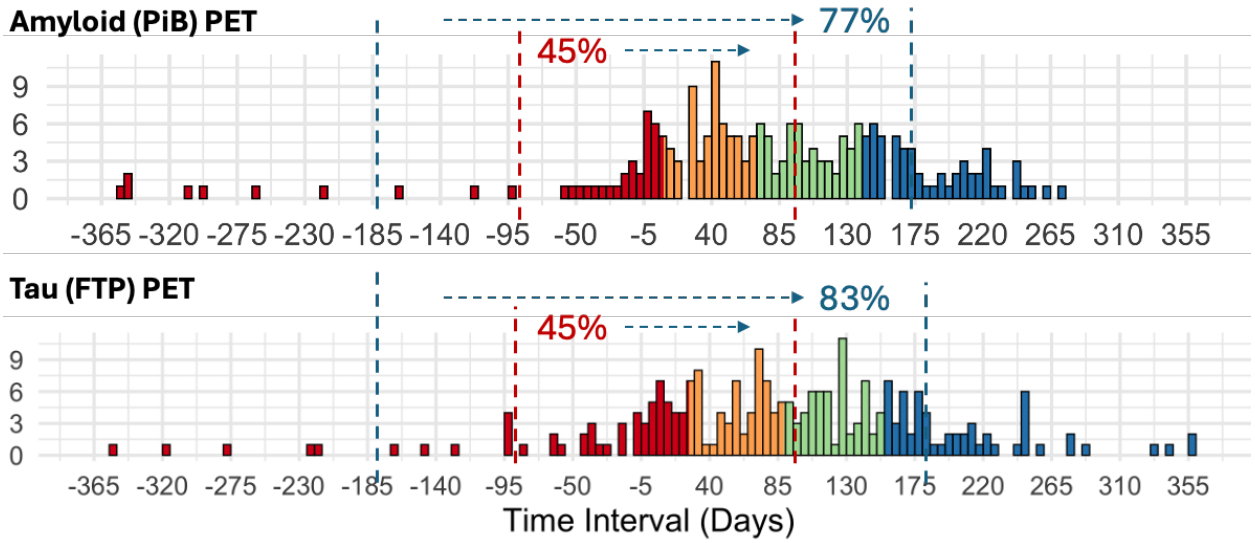


***Note***: Amyloid and tau PET scans were all within a year from MRI; the majority of scan intervals were at or within three-month, followed by six-month target ranges and had no impact on analyses. Colors represent quartiles.

**Figure S2.** ARTS scores by stratified groups

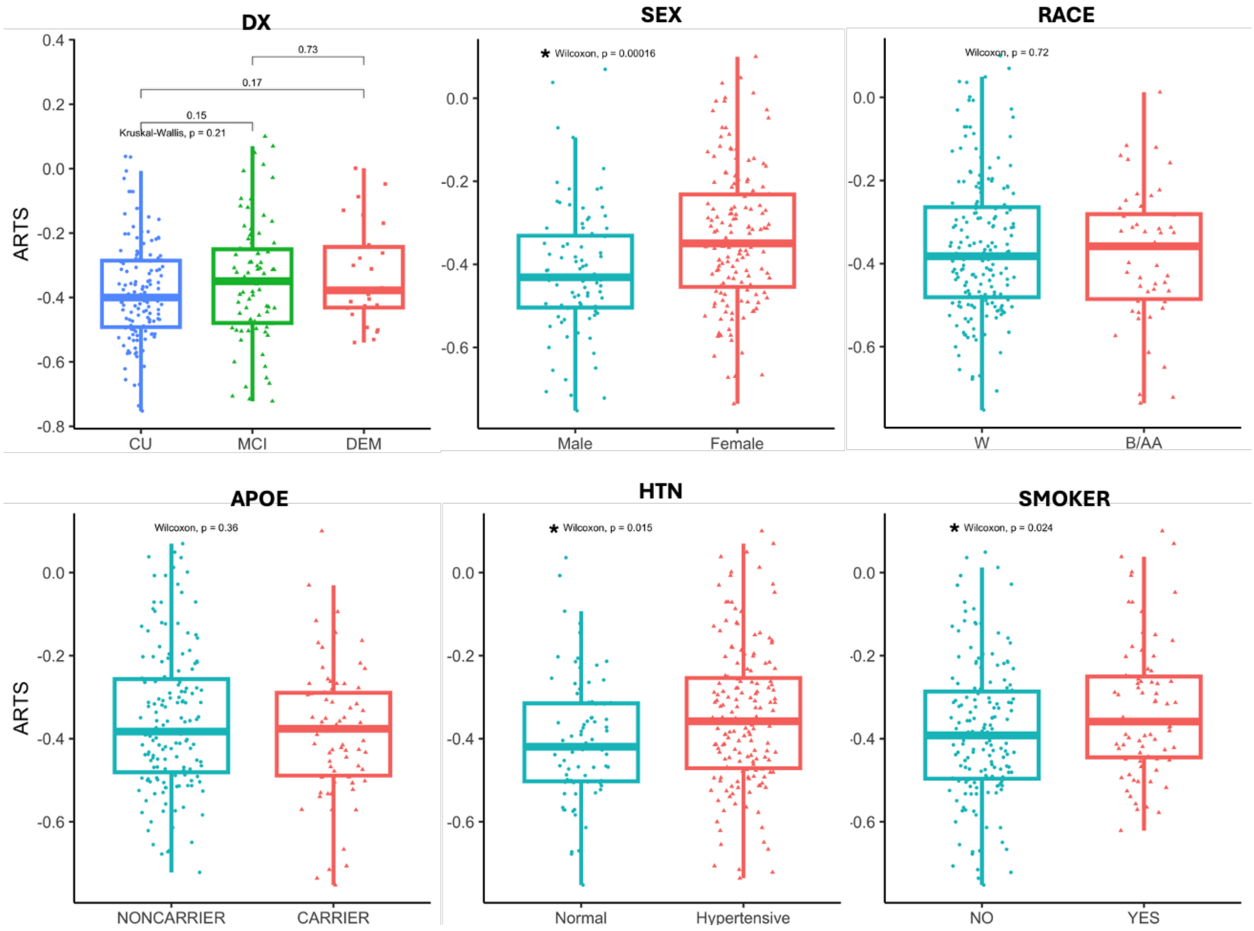


***Note***: ARTS Scores were higher in females compared to males, and participants with hypertension or a history of smoking. Abbreviations: ARTS = Arteriolosclerosis; DX = Cognitive Status; CU = Cognitively Unimpaired; MCI = Mild Cognitive Impairment; DEM = Dementia; HTN = Hypertension

**Figure S3.** Scatterplots of unadjusted correlations between ARTS scores and age at MRI

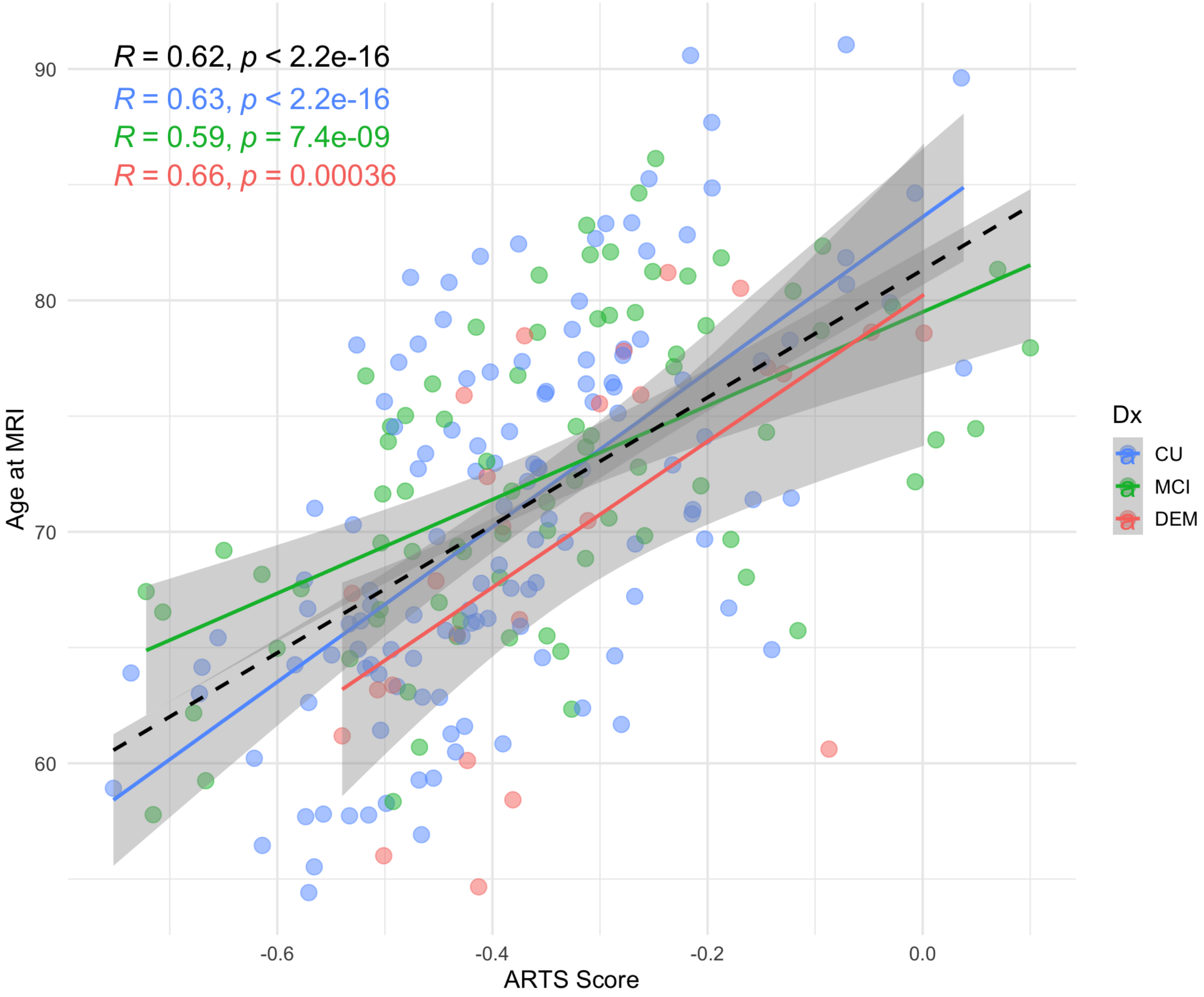


***Note***: Associations between ARTS and age at MRI. Associations between ARTS and age at MRI for the full sample are presented in black and by diagnosis (CU=Blue; MCI=Green; DEM=Red). Abbreviations: ARTS = Arteriolosclerosis; CU = Cognitively Unimpaired; MCI = Mild Cognitive Impairment; DEM = Dementia

**Figure S4.** Unadjusted spearman correlations

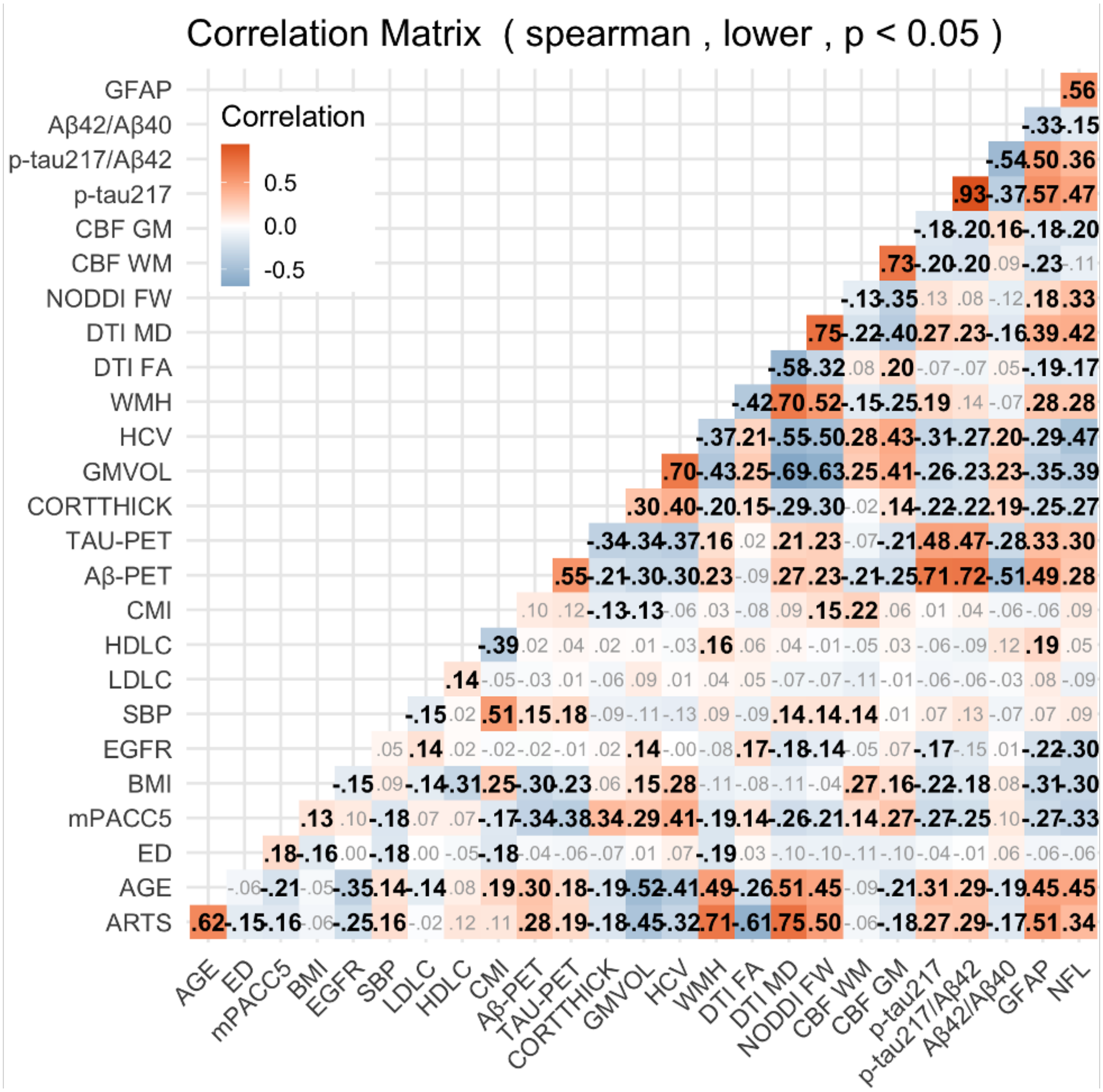


***Note***: Associations between ARTS and all primary variables of interest assessed. Values represent unadjusted Spearman correlation coefficients Significant values ($p < .05$) are displayed in bold black text. Abbreviations: ARTS = Arteriolosclerosis; ED = Education Level; BMI = Body Mass Index; EGFR = Estimated Glomerular Filtration Rate; SBP = Systolic Blood Pressure; LDLC = Low-Density Lipoprotein Cholesterol; HDLC = High-Density Lipoprotein Cholesterol; CMI = Cardiometabolic Index; CORTTHICK = Meta-Temporal Cortical Thickness; GMVOL = ICV Adjusted Total Gray Matter Volume; HCV = Hippocampal Volume; WMH = White Matter Hyperintensity Volume (ICV-Adjusted, Log-Transformed); FA = Fractional Anisotropy; FW = Free Water; CBF = Cerebral Blood Flow; NFL = Neurofilament Light; GFAP = Glial Acidic Fibrillary Acid

Figure S5. Biomarker outcomes stratified by hypertensive status

A. Neuroimaging Biomarkers

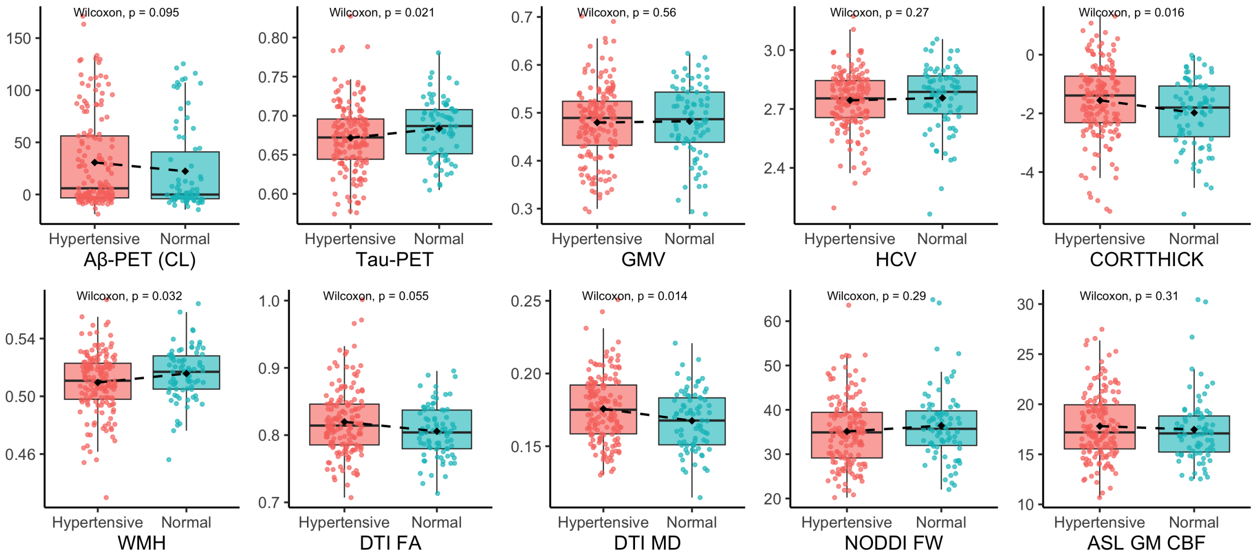


B. Plasma Biomarkers

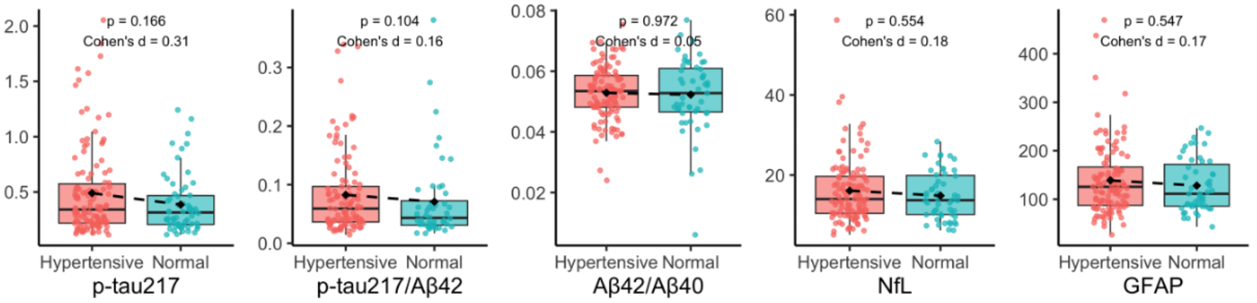

**Figure S6.** Scatterplots of unadjusted correlations between ARTS scores and neuroimaging biomarkers

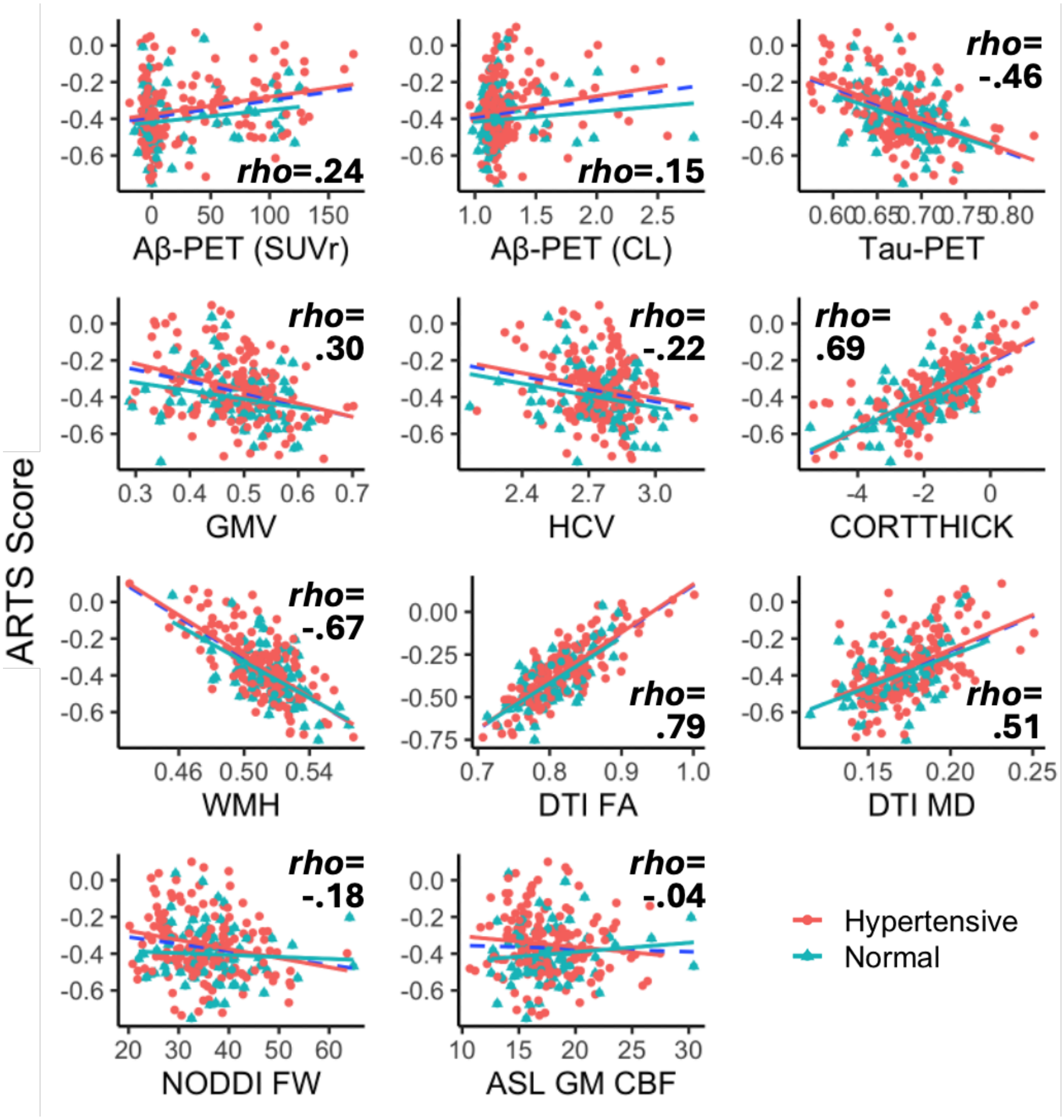


*Note:* Unadjusted correlations between ARTS scores and neuroimaging biomarkers and Spearman correlation coefficients are displayed for the full sample (blue dotted line). Associations are also plotted by hypertension status (Hypertensive = Red; Normal = Teal) for comparison. *Abbreviations*: ARTS = Arteriosclerosis; Aβ = Amyloid Beta; SUVr = Standardized Uptake Volume Ratio; CL = Centiloids; GMV = Total Gray Matter Brain Volume Adjusted for Intracranial Volume; HCV = Hippocampal Volume Adjusted for Intracranial Volume; CORTTHICK = Cortical Thickness; WMH = Log-Transformed White Matter Hyperintensity Volume Adjusted for Intracranial Volume; DTI FA = Diffusion Tensor Imaging Fractional Anisotropy; DTI MD = Diffusion Tensor Imaging Mean Diffusivity; NODDI FW = Neurite Orientation Diffusion Index Free Water; CBF WM = Cerebral Blood Flow in White Matter; CBF GM = Cerebral Blood Flow in Gray Matter

**Figure S7.** Scatterplots of unadjusted correlations between ARTS scores and plasma biomarkers

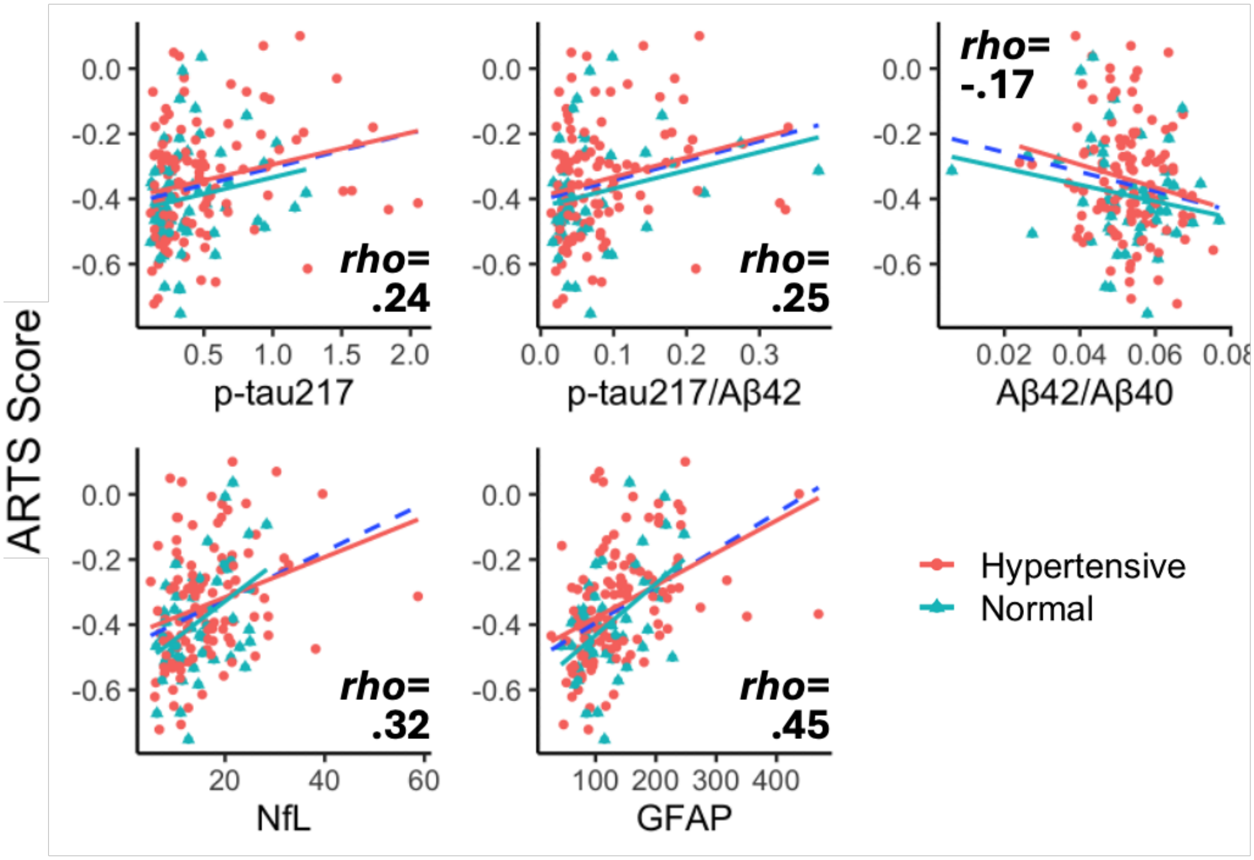


*Note:* Unadjusted correlations between ARTS scores and plasma biomarkers and Spearman correlation coefficients are displayed for the full sample (blue dotted line). Associations are also plotted by hypertension status (Hypertensive = Red; Normal = Teal) for comparison. *Abbreviations*: ARTS = Arteriosclerosis; Aβ = Amyloid Beta; NfL = Neurofilament Light Chain; GFAP = Glial Acid Fibrillary Acid Protein

**Figure S8.** Post-hoc effect modification analyses

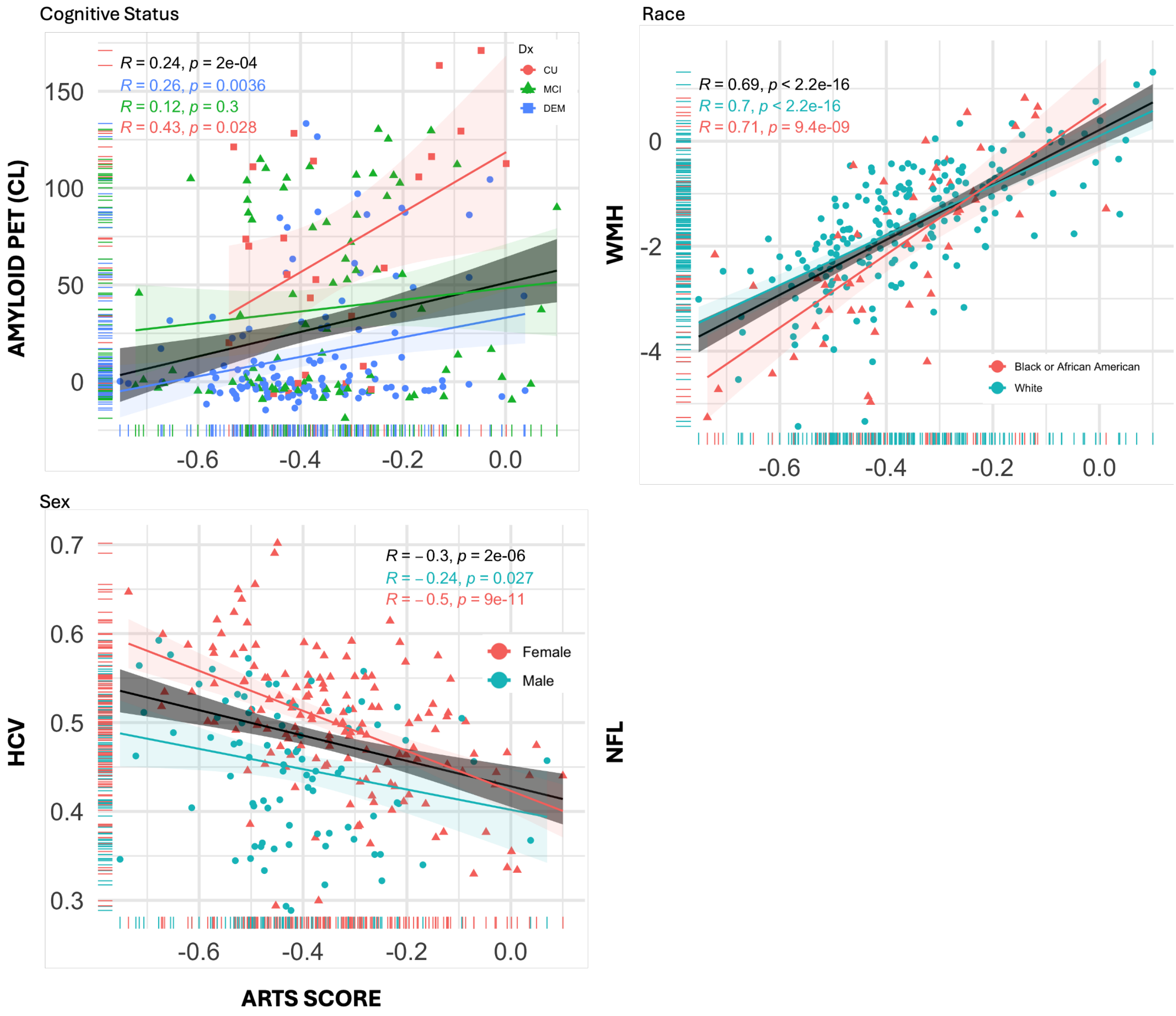


*Note*: Unadjusted scatterplots showing associations between ARTS scores and neuroimaging biomarkers that were modified by diagnostic status, sex, and race respectively. *Abbreviations*: ARTS = Arteriolosclerosis; HCV = Hippocampal Volume; WMH = White Matter Hyperintensity Volume (ICV-Adjusted, Log-Transformed)

**Table S1.** Sample characteristics across analytic and full WFADRC samples

| | N | Overall<br>N = 814[1] | EXCLUDED<br>N = 576[1] | INCLUDED<br>N = 238[1] | p[2] |
|---|---|---|---|---|---|
| **DX** | 814 | | | | .120 |
| **CU** | | 406 (50%) | 274 (48%) | 132 (55%) | |
| **MCI** | | 303 (37%) | 223 (39%) | 80 (34%) | |
| **DEM** | | 105 (13%) | 79 (14%) | 26 (11%) | |
| **AGE** | 814 | 70.67 (8.06) | 70.44 (8.18) | 71.21 (7.73) | .200 |
| **FEMALE** | 814 | 547 (67%) | 397 (69%) | 150 (63%) | .100 |
| **RACE** | 814 | | | | **.033** |
| **American Indian or Alaska Native** | | 4 (0.5%) | 3 (0.5%) | 1 (0.4%) | |
| **Asian** | | 8 (1.0%) | 7 (1.2%) | 1 (0.4%) | |
| **Black or African American** | | 218 (27%) | 168 (29%) | 50 (21%) | |
| **Native Hawaiian or Other Pacific Islander** | | 1 (0.1%) | 0 (0%) | 1 (0.4%) | |
| **White** | | 583 (72%) | 398 (69%) | 185 (78%) | |
| **EDUCATION** | 814 | 15.78 (2.52) | 15.73 (2.50) | 15.91 (2.57) | .400 |
| **SMOKER** | 813 | 258 (32%) | 182 (32%) | 76 (32%) | >0.9 |
| ***APOE*-ε4+** | 770 | 276 (36%) | 202 (37%) | 74 (32%) | .200 |
| **HTN** | 814 | 536 (66%) | 377 (65%) | 159 (67%) | .700 |
| **SYSBP** | 809 | 134.12 (19.53) | 133.62 (18.74) | 135.34 (21.32) | .150 |
| **IGT** | 795 | 476 (60%) | 335 (60%) | 141 (59%) | .800 |
| **BMI** | 800 | 27.86 (6.00) | 27.88 (5.83) | 27.80 (6.40) | .900 |
| **EGFR** | 652 | 76.88 (15.56) | 76.44 (15.93) | 77.69 (14.86) | .500 |

**Note**: Sample characteristics of the analytical subset of participants adjudicated as cognitively unimpaired, MCI, or dementia with ARTS, MRI, and PET data was overall highly comparable to the full WFADRC cohort. The analytic sample with all neuroimaging data had a relatively higher number of White participants similar to previous reports.[3] *Abbreviations*: CU = Cognitively Unimpaired; MCI = Mild Cognitive Impairment; DEM = Dementia; HTN = Hypertension; SYSBP = Systolic Blood Pressure; IGT = Impaired Glucose Tolerance; BMI = Body Mass Index; eGFR = Estimated Glomerular Filtration Rate

**Table S2:** Association of ARTS with amyloid and tau PET using GAMMA regression

| *Gamma** | Model 1 (Unadjusted) | | | | | | Model 2 (Adjusted) | | | | | |
|---|---|---|---|---|---|---|---|---|---|---|---|---|
| | ***n*** | **R2** | ***β*** | ***SE*** | ***p*** | ***q*** | ***n*** | **R2** | ***β*** | ***SE*** | ***p*** | ***Q*** |
| **Aβ (SUVr)** | *234* | *.08* | 0.51 | 0.12 | **<.001** | **<.001** | *225* | .26 | 0.38 | 0.15 | **.010** | **.039** |
| **TAU (SUVr)** | 238 | .03 | 0.19 | 0.08 | **.021** | **.034** | 229 | .13 | 0.33 | 0.10 | **.001** | **.017** |

** Centiloids (CL) excluded - Poisson regression requires non-negative values*

*Note*: Variables were centered and scaled (z-transformed) prior to model generation. All models were fit using robust standard errors. Model 1 was unadjusted. Model 2 was adjusted for age, sex, race, education, APOE, and the interval between PET and MRI scan acquisition. Abbreviations: R2 = Coefficient of Determination (% variance explained); SE = Standard Error; SUVr = Standard-Uptake Value Ratio

**Table S3**. Association of ARTS with amyloid and tau PET positivity

| **A. Aβ-PET+** | **Model 1 (Unadjusted)** | | | | | | **Model 2 (Adjusted)** | | | | | |
|---|---|---|---|---|---|---|---|---|---|---|---|---|
| | *n* | **R2** | ***OR*** | ***SE*** | ***p*** | ***q*** | ***n*** | **R2** | ***OR*** | ***SE*** | ***p*** | ***q*** |
| ***VISREAD*** | *238* | 0.05 | 1.60 | 0.23 | **.001** | **.003** | 229 | 0.23 | 1.36 | 0.27 | .124 | .135 |
| ***SUVr ≧1.21*** | *234* | 0.05 | 1.58 | 0.22 | **.001** | **.003** | 225 | 0.23 | 1.28 | 0.26 | .214 | .214 |
| ***CL ≧12.2*** | *233* | 0.05 | 1.60 | 0.23 | **.001** | **.003** | 225 | 0.23 | 1.40 | 0.28 | .098 | .118 |
| ***CL ≧24*** | *233* | 0.05 | 1.64 | 0.24 | **.001** | **.003** | 225 | 0.18 | 1.42 | 0.29 | .080 | .107 |
| ***B. TAU-PET+*** | **Model 1 (Unadjusted)** | | | | | | **Model 2 (Adjusted)** | | | | | |
| | ***n*** | **R2** | ***OR*** | ***SE*** | ***p*** | ***q*** | ***n*** | **R2** | ***OR*** | ***SE*** | ***p*** | **Q** |
| ***VISREAD*** | *238* | 0.02 | 1.41 | 0.24 | **.040** | .060 | 229 | 0.12 | 1.92 | 0.53 | **.018** | **.036** |
| ***SUVr ≧1.21*** | *238* | 0.03 | 1.46 | 0.20 | **.006** | **.014** | 229 | 0.13 | 1.54 | 0.32 | **.039** | .060 |

*Note*: Variables were standardized (centered and scaled [z-transformed]) prior to model generation. All models were fit using robust standard errors. Model 1 was unadjusted. Model 2 was adjusted for age, sex, race, education, APOE, and the interval between PET and MRI scan acquisition. Abbreviations: R2 = Coefficient of Determination (% variance explained); OR = Odd Ratio; SE = Standard Error; q = FDR-corrected p-values; VISREAD = Visual Read; SUVr = Standard-Uptake Value Ratio; CL = Centiloids

**Table S4**. Significant covariates from regression models assessing associations between ARTS and imaging and plasma biomarkers

| | COVARIATES ( X = SIGNIFICANT BETA ) | | | | | | | |
|---|---|---|---|---|---|---|---|---|
| **OUTCOMES** | **AGE** | **SEX** | **RACE** | **EDUC** | **APOE** | **INT** | **BMI** | **EGFR** |
| **Aβ-PET** | | | x | | x | | -- | -- |
| **TAU-PET** | | | x | | x | | -- | -- |
| **GMV** | x | x | x | | | | -- | -- |
| **HCV** | x | x | x | | | | -- | -- |
| **CORTTHICK** | | | | | | | -- | -- |
| **WMH** | | x | | x | | | -- | -- |
| **DTI FA** | x | x | | | | | -- | -- |
| **DTI MD** | | x | x | | | | -- | -- |
| **NODDI FW** | | x | | | | | -- | -- |
| **CBF WM** | | x | x | | x | | -- | -- |
| **CBF GM** | | x | x | | x | | -- | -- |
| **p-tau217** | | | x | | x | | x | x |
| **p-tau217/Aβ42** | | | | | x | | x | |
| **Aβ42/40** | x | | | | x | | | |
| **NfL** | x | | x | | x | | x | x |
| **GFAP** | x | | | | x | | x | x |

*Note*: Table showing significant covariates ($p<.05$) from adjusted regression models assessing associations between ARTS and imaging and plasma biomarkers. Model 2 was adjusted for age, sex, race, education, APOE, and the interval between biomarker acquisition and MRI, as well as BMI and eGFR for plasma models. *Abbreviations*: CU = Cognitively Unimpaired; MCI = Mild Cognitive Impairment; DEM = Dementia; ARTS = Arteriolosclerosis Score; HTN = Hypertension; SYSBP = Systolic Blood Pressure; IGT = Impaired Glucose Tolerance; Aβ = Amyloid Beta; GMV = Total Gray Matter Brain Volume Adjusted for Intracranial Volume; HCV = Hippocampal Volume Adjusted for Intracranial Volume; CORTTHICK = Cortical Thickness; WMH = Log-Transformed White Matter Hyperintensity Volume Adjusted for Intracranial Volume; DTI FA = Diffusion Tensor Imaging Fractional Anisotropy; DTI MD = Diffusion Tensor Imaging Mean Diffusivity; NODDI FW = Neurite Orientation Diffusion Index Free Water; CBF WM = Cerebral Blood Flow in White Matter; CBF GM = Cerebral Blood Flow in Gray Matter; NfL = Neurofilament Light Chain; GFAP = Glial Acid Fibrillary Acid Protein; EDUC = Education; INT = Interval Between MRI Scan or Blood Draw and MRI; BMI = Body Mass Index; eGFR = Estimated Glomerular Filtration Rate

**Table S5.** Sample characteristics stratified by canonical ATN status

| | N | Overall<br>N = 238[1] | A-T-N-<br>N = 84[1] | A+T-N-<br>N = 25[1] | A+T+N-<br>N = 26[1] | A+T+N+<br>N = 39[1] | OTHER<br>N = 64[1] | p[2] |
|---|---|---|---|---|---|---|---|---|
| **DX** | 238 | | | | | | | **<.001** |
| **CU** | | 132 (55%) | 66 (79%) | 19 (76%) | 9 (35%) | 6 (15%) | 32 (50%) | |
| **MCI** | | 80 (34%) | 15 (18%) | 4 (16%) | 14 (54%) | 21 (54%) | 26 (41%) | |
| **DEM** | | 26 (11%) | 3 (3.6%) | 2 (8.0%) | 3 (12%) | 12 (31%) | 6 (9.4%) | |
| **AGE (MRI)** | 238 | 71 (8) | 68 (7) | 72 (7) | 71 (8) | 75 (8) | 73 (7) | **<.001** |
| **SEX:FEMALE** | 238 | 150 (63%) | 60 (71%) | 18 (72%) | 15 (58%) | 21 (54%) | 36 (56%) | .200 |
| **RACE** | 238 | | | | | | | **.035** |
| **White** | | 185 (78%) | 60 (71%) | 19 (76%) | 21 (81%) | 38 (97%) | 47 (73%) | |
| **AA/BLACK** | | 50 (21%) | 22 (26%) | 6 (24%) | 5 (19%) | 1 (2.6%) | 16 (25%) | |
| **AI/AN** | | 1 (0.4%) | 1 (1.2%) | 0 (0%) | 0 (0%) | 0 (0%) | 0 (0%) | |
| **NH/PI** | | 1 (0.4%) | 0 (0%) | 0 (0%) | 0 (0%) | 0 (0%) | 1 (1.6%) | |
| **Asian** | | 1 (0.4%) | 1 (1.2%) | 0 (0%) | 0 (0%) | 0 (0%) | 0 (0%) | |
| **EDUCATION** | 238 | 15.91 (2.57) | 16.04 (2.50) | 16.24 (2.54) | 16.00 (2.70) | 15.56 (2.78) | 15.78 (2.56) | .800 |
| **mPACC5** | 236 | -0.59 (1.13) | -0.10 (0.84) | -0.21 (0.93) | -0.86 (1.16) | -1.78 (1.24) | -0.58 (0.93) | **<.001** |
| **SMOKER** | 238 | 76 (32%) | 26 (31%) | 7 (28%) | 11 (42%) | 14 (36%) | 18 (28%) | .700 |
| ***APOE*-ε4+** | 229 | 74 (32%) | 16 (20%) | 8 (33%) | 14 (56%) | 20 (53%) | 16 (27%) | **<.001** |
| **ARTS** | 238 | -0.37 (0.17) | -0.43 (0.15) | -0.36 (0.16) | -0.32 (0.16) | -0.31 (0.18) | -0.35 (0.16) | **<.001** |
| **HTN** | 238 | 159 (67%) | 46 (55%) | 20 (80%) | 23 (88%) | 27 (69%) | 43 (67%) | **.011** |
| **SYSBP** | 238 | 135 (21) | 130 (22) | 138 (18) | 140 (17) | 137 (23) | 138 (22) | .080 |
| **IGT** | 238 | 141 (59%) | 47 (56%) | 19 (76%) | 16 (62%) | 18 (46%) | 41 (64%) | .200 |
| **BMI** | 238 | 27.8 (6.4) | 29.5 (6.1) | 27.9 (7.4) | 27.3 (5.8) | 23.0 (5.6) | 28.6 (5.7) | **<.001** |
| **eGFR** | 230 | 78 (15) | 79 (14) | 76 (18) | 76 (13) | 81 (14) | 75 (15) | .200 |
| **HX TBI** | 238 | 6 (2.5%) | 2 (2.4%) | 1 (4.0%) | 1 (3.8%) | 0 (0%) | 2 (3.1%) | .700 |
| **HX SLEEP APNEA** | 238 | 67 (28%) | 24 (29%) | 6 (24%) | 10 (38%) | 8 (21%) | 19 (30%) | .600 |
| | | | | **PET IMAGING** | | | | |
| **Aβ+ (Read)** | 238 | 102 (43%) | 0 (0%) | 25 (100%) | 26 (100%) | 39 (100%) | 12 (19%) | **<.001** |
| **Aβ+ (CL > 24)** | 233 | 85 (36%) | 0 (0%) | 18 (75%) | 25 (96%) | 35 (90%) | 7 (11%) | **<.001** |
| **Aβ-PET (SUVr)** | 234 | 1.40 (0.45) | 1.09 (0.07) | 1.60 (0.37) | 1.82 (0.41) | 2.01 (0.39) | 1.17 (0.22) | **<.001** |
| **Aβ-PET (CL)** | 233 | 28 (44) | -2 (5) | 45 (33) | 72 (38) | 87 (39) | 5 (22) | **<.001** |
| **Aβ-PET-MRI-INT** | 238 | 24 (222) | 7 (233) | 48 (176) | 42 (188) | 42 (218) | 17 (241) | >.900 |
| **TAU-PET+ (SUVr)** | 238 | 97 (41%) | 0 (0%) | 0 (0%) | 26 (100%) | 39 (100%) | 32 (50%) | **<.001** |
| **TAU PET (SUVr)** | 238 | 1.27 (0.27) | 1.13 (0.05) | 1.14 (0.04) | 1.45 (0.36) | 1.63 (0.36) | 1.20 (0.08) | **<.001** |
| **TAU-PET-MRI-INT** | 238 | 86 (107) | 77 (118) | 93 (107) | 82 (113) | 100 (87) | 88 (104) | .900 |
| | | | | **NEUROIMAGING** | | | | |
| **GMV** | 238 | 0.68 (0.04) | 0.70 (0.04) | 0.69 (0.03) | 0.67 (0.04) | 0.64 (0.03) | 0.66 (0.05) | **<.001** |
| **HCV** | 238 | 0.48 (0.08) | 0.53 (0.05) | 0.52 (0.04) | 0.51 (0.05) | 0.39 (0.04) | 0.44 (0.08) | **<.001** |
| **CORTTHICK** | 238 | 2.75 (0.15) | 2.80 (0.13) | 2.79 (0.14) | 2.76 (0.12) | 2.64 (0.16) | 2.72 (0.16) | **<.001** |
| **WMH (log/ICV)** | 238 | -1.70 (1.26) | -2.08 (1.27) | -1.69 (1.17) | -1.47 (1.44) | -1.08 (0.92) | -1.66 (1.24) | **<.001** |
| **DTI FA** | 238 | 0.512 (0.020) | 0.515 (0.019) | 0.509 (0.025) | 0.512 (0.017) | 0.510 (0.022) | 0.510 (0.020) | .600 |
| **DTI MD** | 238 | 0.82 (0.05) | 0.80 (0.04) | 0.81 (0.04) | 0.82 (0.05) | 0.84 (0.04) | 0.82 (0.05) | **<.001** |
| **NODDI FW** | 237 | 0.173 (0.022) | 0.164 (0.020) | 0.169 (0.022) | 0.177 (0.021) | 0.185 (0.023) | 0.177 (0.021) | **<0001** |
| **ASL WM CBF** | 234 | 17.7 (3.3) | 18.7 (3.5) | 17.3 (3.4) | 16.6 (2.2) | 16.6 (3.0) | 17.6 (3.3) | **.004** |
| **ASL GM CBF** | 234 | 36 (8) | 39 (8) | 36 (8) | 32 (4) | 32 (6) | 34 (8) | **<.001** |
| | | | | **PLASMA** | | | | |
| **p-tau217** | 184 | 0.45 (0.36) | 0.25 (0.13) | 0.55 (0.34) | 0.71 (0.46) | 0.89 (0.43) | 0.33 (0.17) | **<.001** |
| **p-tau217/Aβ42** | 158 | 0.08 (0.07) | 0.04 (0.02) | 0.10 (0.07) | 0.13 (0.07) | 0.16 (0.08) | 0.06 (0.06) | **<0.001** |
| **Aβ42/40** | 166 | 0.053 (0.010) | 0.057 (0.008) | 0.048 (0.008) | 0.050 (0.006) | 0.047 (0.007) | 0.052 (0.012) | **<.001** |

| | | | | | | | | |
|---|---|---|---|---|---|---|---|---|
| **NfL** | 166 | 16 (7) | 12 (5) | 15 (7) | 16 (6) | 20 (6) | 18 (10) | **<.001** |
| **GFAP** | 166 | 136 (68) | 102 (39) | 142 (54) | 181 (105) | 184 (77) | 132 (57) | **<.001** |

[1] n (%); Mean (SD); [2] Kruskal-Wallis rank sum test; Fisher's exact test

*Note*. Sample characteristics stratified by ATN status are provided in Table S2. *Abbreviations*: CU = Cognitively Unimpaired; MCI = Mild Cognitive Impairment; DEM = Dementia; ARTS = Arteriolosclerosis Score; HTN = Hypertension; SYSBP = Systolic Blood Pressure; IGT = Impaired Glucose Tolerance; BMI = Body Mass Index; eGFR = Estimated Glomerular Filtration Rate; HX TBI = History of Traumatic Brain Injury; Aβ = Amyloid Beta; SUVr = Standardized Uptake Volume Ratio; CL = Centiloids; GMV = Total Gray Matter Brain Volume Adjusted for Intracranial Volume; HCV = Hippocampal Volume Adjusted for Intracranial Volume; CORTTHICK = Cortical Thickness; WMH = Log-Transformed White Matter Hyperintensity Volume Adjusted for Intracranial Volume; DTI FA = Diffusion Tensor Imaging Fractional Anisotropy; DTI MD = Diffusion Tensor Imaging Mean Diffusivity; NODDI FW = Neurite Orientation Diffusion Index Free Water; CBF WM = Cerebral Blood Flow in White Matter; CBF GM = Cerebral Blood Flow in Gray Matter; NfL = Neurofilament Light Chain; GFAP = Glial Acid Fibrillary Acid Protein; PTAU-MRI-INT = Interval Between Blood Draw and MRI for p-tau217; NCRAD -MRI-INT = Interval Between Blood Draw and MRI for NfL and GFAP

**Table S6.** Sample characteristics stratified by ATN group

| | N | Overall N = 238[1] | A-T-N- N = 84[1] | A-T-N+ N = 20[1] | A-T+N- N = 23[1] | A-T+N+ N = 9[1] | A+T-N- N = 25[1] | A+T-N+ N = 12[1] | A+T+N- N = 26[1] | A+T+N+ N = 39[1] |
|---|---|---|---|---|---|---|---|---|---|---|
| **DX** | 238 | | | | | | | | | |
| **CU** | | 132 (55%) | 66 (79%) | 9 (45%) | 14 (61%) | 4 (44%) | 19 (76%) | 5 (42%) | 9 (35%) | 6 (15%) |
| **MCI** | | 80 (34%) | 15 (18%) | 8 (40%) | 9 (39%) | 5 (56%) | 4 (16%) | 4 (33%) | 14 (54%) | 21 (54%) |
| **DEM** | | 26 (11%) | 3 (3.6%) | 3 (15%) | 0 (0%) | 0 (0%) | 2 (8.0%) | 3 (25%) | 3 (12%) | 12 (31%) |
| **AGE (MRI)** | 238 | 71 (8) | 68 (7) | 71 (7) | 72 (7) | 73 (4) | 72 (7) | 77 (8) | 71 (8) | 75 (8) |
| **SEX:FEMALE** | 238 | 150 (63%) | 60 (71%) | 9 (45%) | 19 (83%) | 3 (33%) | 18 (72%) | 5 (42%) | 15 (58%) | 21 (54%) |
| **RACE** | 238 | | | | | | | | | |
| **White** | | 185 (78%) | 60 (71%) | 16 (80%) | 10 (43%) | 9 (100%) | 19 (76%) | 12(100%) | 21 (81%) | 38 (97%) |
| **AA/BLACK** | | 50 (21%) | 22 (26%) | 4 (20%) | 12 (52%) | 0 (0%) | 6 (24%) | 0 (0%) | 5 (19%) | 1 (2.6%) |
| **AI/AN** | | 1 (0.4%) | 1 (1.2%) | 0 (0%) | 0 (0%) | 0 (0%) | 0 (0%) | 0 (0%) | 0 (0%) | 0 (0%) |
| **NH/PI** | | 1 (0.4%) | 0 (0%) | 0 (0%) | 1 (4.3%) | 0 (0%) | 0 (0%) | 0 (0%) | 0 (0%) | 0 (0%) |
| **Asian** | | 1 (0.4%) | 1 (1.2%) | 0 (0%) | 0 (0%) | 0 (0%) | 0 (0%) | 0 (0%) | 0 (0%) | 0 (0%) |
| **EDUCATION** | 238 | 15.91 (2.57) | 16.04 (2.50) | 15.45 (2.37) | 15.78 (2.54) | 16.56 (3.21) | 16.24 (2.54) | 15.75 (2.60) | 16.00 (2.70) | 15.56 (2.78) |
| **mPACC5** | 236 | -0.59 (1.13) | -0.10 (0.84) | -0.78 (1.09) | -0.36 (0.69) | -0.46 (1.05) | -0.21 (0.93) | -0.76 (0.96) | -0.86 (1.16) | -1.78 (1.24) |
| **SMOKER** | 238 | 76 (32%) | 26 (31%) | 6 (30%) | 4 (17%) | 4 (44%) | 7 (28%) | 4 (33%) | 11 (42%) | 14 (36%) |
| ***APOE*-ε4+** | 229 | 74 (32%) | 16 (20%) | 3 (17%) | 5 (24%) | 2 (22%) | 8 (33%) | 6 (50%) | 14 (56%) | 20 (53%) |
| **ARTS** | 238 | -0.37 (0.17) | -0.43 (0.15) | -0.33 (0.17) | -0.39 (0.14) | -0.31 (0.20) | -0.36 (0.16) | -0.31 (0.11) | -0.32 (0.16) | -0.31 (0.18) |
| **HTN** | 238 | 159 (67%) | 46 (55%) | 10 (50%) | 18 (78%) | 7 (78%) | 20 (80%) | 8 (67%) | 23 (88%) | 27 (69%) |
| **SYSBP** | 238 | 135 (21) | 130 (22) | 137 (20) | 138 (22) | 146 (19) | 138 (18) | 136 (27) | 140 (17) | 137 (23) |
| **PRE/DIABETES** | 238 | 141 (59%) | 47 (56%) | 9 (45%) | 19 (83%) | 6 (67%) | 19 (76%) | 7 (58%) | 16 (62%) | 18 (46%) |
| **BMI** | 238 | 27.8 (6.4) | 29.5 (6.1) | 27.0 (4.4) | 30.8 (6.1) | 27.5 (6.6) | 27.9 (7.4) | 27.8 (5.3) | 27.3 (5.8) | 23.0 (5.6) |
| **eGFR** | 230 | 78 (15) | 79 (14) | 82 (15) | 70 (14) | 81 (14) | 76 (18) | 68 (12) | 76 (13) | 81 (14) |
| **HX TBI** | 238 | 6 (2.5%) | 2 (2.4%) | 1 (5.0%) | 0 (0%) | 0 (0%) | 1 (4.0%) | 1 (8.3%) | 1 (3.8%) | 0 (0%) |
| **HX SLEEP APNEA** | 238 | 67 (28%) | 24 (29%) | 4 (20%) | 9 (39%) | 5 (56%) | 6 (24%) | 1 (8.3%) | 10 (38%) | 8 (21%) |
| | | | | | **PET IMAGING** | | | | | |
| **Aβ+ (Read)** | 238 | 102 (43%) | 0 (0%) | 0 (0%) | 0 (0%) | 0 (0%) | 25 (100%) | 12 (100%) | 26 (100%) | 39 (100%) |
| **Aβ+ (CL > 24)** | 233 | 85 (36%) | 0 (0%) | 0 (0%) | 0 (0%) | 0 (0%) | 18 (75%) | 7 (58%) | 25 (96%) | 35 (90%) |
| **Aβ-PET (SUVr)** | 234 | 1.40 (0.45) | 1.09 (0.07) | 1.06 (0.08) | 1.09 (0.08) | 1.13 (0.05) | 1.60 (0.37) | 1.51 (0.31) | 1.82 (0.41) | 2.01 (0.39) |
| **Aβ-PET (CL)** | 233 | 28 (44) | -2 (5) | -3 (6) | -3 (6) | -3 (4) | 45 (33) | 37 (33) | 72 (38) | 87 (39) |
| **Aβ-PET-MRI-INT** | 238 | 24 (222) | 7 (233) | 36 (193) | -20 (316) | 11 (255) | 48 (176) | 63 (121) | 42 (188) | 42 (218) |
| **TAU-PET+ (SUVr)** | 238 | 97 (41%) | 0 (0%) | 0 (0%) | 23 (100%) | 9 (100%) | 0 (0%) | 0 (0%) | 26 (100%) | 39 (100%) |

| | | | | | | | | | | |
|---|---|---|---|---|---|---|---|---|---|---|
| **TAU PET (SUVr)** | 238 | 1.27 (0.27) | 1.13 (0.05) | 1.13 (0.06) | 1.25 (0.05) | 1.28 (0.03) | 1.14 (0.04) | 1.15 (0.04) | 1.45 (0.36) | 1.63 (0.36) |
| **TAU-PET-MRI-INT** | 238 | 86 (107) | 77 (118) | 65 (127) | 100 (106) | 72 (84) | 93 (107) | 112 (65) | 82 (113) | 100 (87) |
| **NEUROIMAGING** | | | | | | | | | | |
| **GMV** | 238 | 0.68 (0.04) | 0.70 (0.04) | 0.65 (0.04) | 0.69 (0.05) | 0.64 (0.02) | 0.69 (0.03) | 0.64 (0.04) | 0.67 (0.04) | 0.64 (0.03) |
| **HCV** | 238 | 0.48 (0.08) | 0.53 (0.05) | 0.40 (0.06) | 0.52 (0.05) | 0.38 (0.04) | 0.52 (0.04) | 0.41 (0.03) | 0.51 (0.05) | 0.39 (0.04) |
| **CORTTHICK** | 238 | 2.75 (0.15) | 2.80 (0.13) | 2.69 (0.16) | 2.76 (0.13) | 2.63 (0.22) | 2.79 (0.14) | 2.78 (0.13) | 2.76 (0.12) | 2.64 (0.16) |
| **WMH (log/ICV)** | 238 | -1.70 (1.26) | -2.08 (1.27) | -1.15 (0.82) | -2.27 (1.40) | -1.61 (1.32) | -1.69 (1.17) | -1.37 (1.03) | -1.47 (1.44) | -1.08 (0.92) |
| **DTI FA** | 238 | 0.512 (0.020) | 0.515 (0.019) | 0.506 (0.019) | 0.517 (0.020) | 0.503 (0.020) | 0.509 (0.025) | 0.507 (0.017) | 0.512 (0.017) | 0.510 (0.022) |
| **DTI MD** | 238 | 0.82 (0.05) | 0.80 (0.04) | 0.84 (0.04) | 0.79 (0.04) | 0.84 (0.06) | 0.81 (0.04) | 0.84 (0.04) | 0.82 (0.05) | 0.84 (0.04) |
| **NODDI FW** | 237 | 0.173 (0.022) | 0.164 (0.020) | 0.185 (0.018) | 0.166 (0.016) | 0.184 (0.020) | 0.169 (0.022) | 0.181 (0.026) | 0.177 (0.021) | 0.185 (0.023) |
| **ASL WM CBF** | 234 | 17.7 (3.3) | 18.7 (3.5) | 16.9 (2.8) | 19.3 (3.6) | 16.1 (4.0) | 17.3 (3.4) | 16.7 (1.8) | 16.6 (2.2) | 16.6 (3.0) |
| **ASL GM CBF** | 234 | 36 (8) | 39 (8) | 33 (8) | 38 (8) | 30 (9) | 36 (8) | 32 (4) | 32 (4) | 32 (6) |
| **PLASMA** | | | | | | | | | | |
| **p-tau217** | 184 | 0.45 (0.36) | 0.25 (0.13) | 0.26 (0.09) | 0.29 (0.13) | 0.29 (0.14) | 0.55 (0.34) | 0.50 (0.23) | 0.71 (0.46) | 0.89 (0.43) |
| **p-tau217/Aβ42** | 158 | 0.08 (0.07) | 0.04 (0.02) | 0.05 (0.02) | 0.08 (0.09) | 0.04 (0.02) | 0.10 (0.07) | 0.09 (0.05) | 0.13 (0.07) | 0.16 (0.08) |
| **Aβ42/40** | 166 | 0.053 (0.010) | 0.057 (0.008) | 0.057 (0.011) | 0.049 (0.014) | 0.059 (0.010) | 0.048 (0.008) | 0.046 (0.009) | 0.050 (0.006) | 0.047 (0.007) |
| **NfL** | 166 | 16 (7) | 12 (5) | 19 (13) | 14 (4) | 20 (11) | 15 (7) | 22 (7) | 16 (6) | 20 (6) |
| **GFAP** | 166 | 136 (68) | 102 (39) | 130 (60) | 120 (63) | 122 (25) | 142 (54) | 158 (55) | 181 (105) | 184 (77) |

[1] n (%); Mean (SD); [2] Kruskal-Wallis rank sum test; Fisher's exact test

*Abbreviations*: CU = Cognitively Unimpaired; MCI = Mild Cognitive Impairment; DEM = Dementia; ARTS = Arteriolosclerosis Score; HTN = Hypertension; SYSBP = Systolic Blood Pressure; IGT = Impaired Glucose Tolerance; BMI = Body Mass Index; eGFR = Estimated Glomerular Filtration Rate; HX TBI = History of Traumatic Brain Injury; Aβ = Amyloid Beta; SUVr = Standardized Uptake Volume Ratio; CL = Centiloids; GMV = Total Gray Matter Brain Volume Adjusted for Intracranial Volume; HCV = Hippocampal Volume Adjusted for Intracranial Volume; CORTTHICK = Cortical Thickness; WMH = Log-Transformed White Matter Hyperintensity Volume Adjusted for Intracranial Volume; DTI FA = Diffusion Tensor Imaging Fractional Anisotropy; DTI MD = Diffusion Tensor Imaging Mean Diffusivity; NODDI FW = Neurite Orientation Diffusion Index Free Water; CBF WM = Cerebral Blood Flow in White Matter; CBF GM = Cerebral Blood Flow in Gray Matter; NfL = Neurofilament Light Chain; GFAP = Glial Acid Fibrillary Acid Protein; PTAU-MRI-INT = Interval Between Blood Draw and MRI for p-tau217; NCRAD -MRI-INT = Interval Between Blood Draw and MRI for NfL and GFAP
